# Plateaus, Rebounds and the Effects of Individual Behaviours in Epidemics

**DOI:** 10.1101/2021.03.26.21254414

**Authors:** Henri Berestycki, Benoît Desjardins, Bruno Heintz, Jean-Marc Oury

## Abstract

Plateaus and rebounds of various epidemiological indicators are widely reported in Covid-19 pandemics studies but have not been explained so far. Here, we address this problem and explain the appearance of these patterns. We start with an empirical study of an original dataset obtained from highly precise measurements of SARS-Cov-2 concentration in wastewater over nine months in several treatment plants around the Thau lagoon in France. Among various features, we observe that the concentration displays plateaus at different dates in various locations but at the same level. In order to understand these facts, we introduce a new mathematical model that takes into account the heterogeneity and the natural variability of individual behaviours. Our model shows that the distribution of risky behaviours appears as the key ingredient for understanding the observed temporal patterns of epidemics.

## 1 Introduction

The onset of plateaus for various indicators of the current outbreak of Covid-19 such as incidence rate or hospitalisations appears to be a rather general feature of its dynamics, along with periods of exponential growth or decay, rebounds etc. Nonetheless, there are few theoretical explanations offered to understand this phenomenon and such plateaus hardly agree with the classical SIR paradigm of epidemics.

We show here that plateaus emerge *intrinsically* in the unfolding of an epidemic. That is, plateaus arise naturally if we take into account two elements: an underlying heterogeneity and a random variability of behaviours in the population. These features are of course more realistic than assuming that the population is perfectly homogeneous with an unwavering behaviour.

To shed light on this mechanism, we propose in this paper a new mathematical model. It takes the form of a system of reaction-diffusion equations, where one variable represents the behaviour of individuals (see Methods section). It is natural to consider that individuals may change their behaviour from one day to the next one. We assume here that individuals’ behaviours move randomly according to Brownian motion among these classes. We show here that such a system that includes heterogeneity and variability of behaviours exhibits a richness of dynamics and in particular gives rise to intrinsic formation of plateaus, shoulders and rebounds.

Up to now, there are two main alternative explanations for the onset of plateaus. The first one is political (1). By managing the epidemic and keeping the exponential growth at bay, without destroying the economy, a plateau appears as some kind of optimal compromise under constraint. Another approach appears in a very recent work of Weitz et al. (2). It argues that plateaus are caused by change of behaviours due to awareness of fatalities and fatigue of the public facing regulatory mobility restrictions. In recent works, Arthur et al. (3) and Radicchi et al. (4) proposed models in the same spirit. More detailed discussion of related literature is provided below.

We then introduce and discuss the mathematical model. We illustrate the types of dynamics that this model gives rise to by numerical simulations. These shed light on the key role of behavioural variability to obtain plateaus, shoulders and rebounds. There, we also provide a simulation to describe the effect of introducing a second variant of the virus that yields a higher secondary epidemic peak.

To discuss the validity of our approach, we rely on observations stemming from a series of measurements, carried weekly over an extended period in the Thau lagoon area in South of France. These strikingly reveal the formation of plateaus, in some cases after a “shoulder” pattern. These data do not include the effect of variants or of rebounds.

Next, we report on the calibration of our model on the data of the Thau lagoon. It yields a remarkable fit. We further discuss our model in more detail in the light of the measurements in the Discussion section below. We also show that this model also generates rebounds.

## 2 Brief review of literature

Numerous papers (5–8) describe the number of daily social contacts as a key variable in the spread of infectious diseases like Covid-19 insofar as it is closely related to the transmission rate. Daily social contacts are usually described in terms of age, gender, income, type of job, household size (9), etc. Parameters that are particularly relevant in the context of viral outbreaks are also studied (10) such as cumulative duration of such contacts, social distance, indoor/outdoor environment, etc. The very fine grain microscopic models (11) aim at identifying such parameters in the most precise way possible. A very recent study by Di Domenico et al. (12) takes up the data of hospitalisations in France for the past six months. Again, these exhibit striking epidemic plateaus since the beginning of 2021. The authors of this paper provide a microscopic insight of the propagation, emphasising the role of two different strains of the virus and the role of public health measures such as the curfew, school closing etc. These approaches are different from ours, as the point of view we adopt here can be seen as “mesoscopic”.

Several earlier works have considered SIR-type systems (Susceptible-Infectious-Removed) with heterogeneity. In particular, Arino et al. (13), and, more recently, Dolbeault and Turinici (14, 15), Magal et al. (16) have studied models with a finite number of different coefficients *β*. These systems are characterised by a discrete set of classes and do not involve variability. Almeida et al. (17) considered the case of continuous classes associated with a multidimensional trait *x* to mathematically study the influence of variability of infectious individuals on the final size of an epidemic. Note that they include a diffusion term of the infectious population while we consider social diffusion of the susceptible.

Weitz et al. (2) recently developed an SEIR-type (Susceptible-Exposed-Infectious-Removed) compartmental model with variable transmission rate coefficient, in which two main competing psychological reactions to the epidemic generate plateaus. Namely, it involves awareness of fatalities and fatigue of the public facing mobility restrictions. In this interesting model, fatigue modulates the transmission rate *β* in the SEIR system by the number of cumulative deaths (and in another version the daily number of deaths). Note that this value itself is an outcome of the model and thus this model is a kind of fixed point formulation. The authors show that their model yields plateaus, “shoulder” like patterns and oscillations for the dynamics of infectious individuals.

This paper of Weitz et al. assumes a homogeneous population: at a given time, all individuals have the same transmission rate. Thus our model is quite different from theirs. The common behaviour simply changes in time by reacting to the outcome of the epidemic and this change reflects in the evolution of the transmission rate. Moreover, we note that the model is calibrated with reported death of various U.S. states and does not involve wastewater concentration measurements. Thus, even though in a different manner from ours, this work also stresses the role of variability for the observed dynamics.

DiMarco et al. (18) have recently proposed a model close in spirit to ours but more complex. Based on kinetic theory, it describes the heterogeneity of individuals in terms of a variable *x* ≥ 0 corresponding to the number of daily social contacts. Their model consists of a system of three SIR equations coupled with Boltzmann or Fokker-Planck type equations. The authors emphasise a collision type term in the resulting Boltzmann equations. This term represents changes of behaviours (that is of the values of *x*) when two individuals meet. Consequently, the present work is quite different from theirs. As a closure of their model, DiMarco et al. (18) formally derive a so called S-SIR model. There, the variability of behaviours involves an explicit dependence of the transmission rates upon the current number of infectious individuals (19). Because of this feature, as a matter of fact, when applied to real data, it is eventually very similar to the approach in Weitz et al. (2)

## Results

### 3 The Thau lagoon data

The measurement campaign concerned four wastewater treatment plants (WWTP) in the Thau lagoon area in France, serving the cities of Sète, Pradel-Marseillan, Frontignan and Mèze. The measurements were obtained by using digital PCR (20) (dPCR) to estimate the concentration of SARS-Cov-2 virus in samples taken weekly from 2020-05-12 to 2021-01-12. We provide further details about the measurement method in the Methods section.

Figure 1 shows the outcomes in a logarithmic scale over a nine months period. We summarise now their main features.

**Fig. 1.**
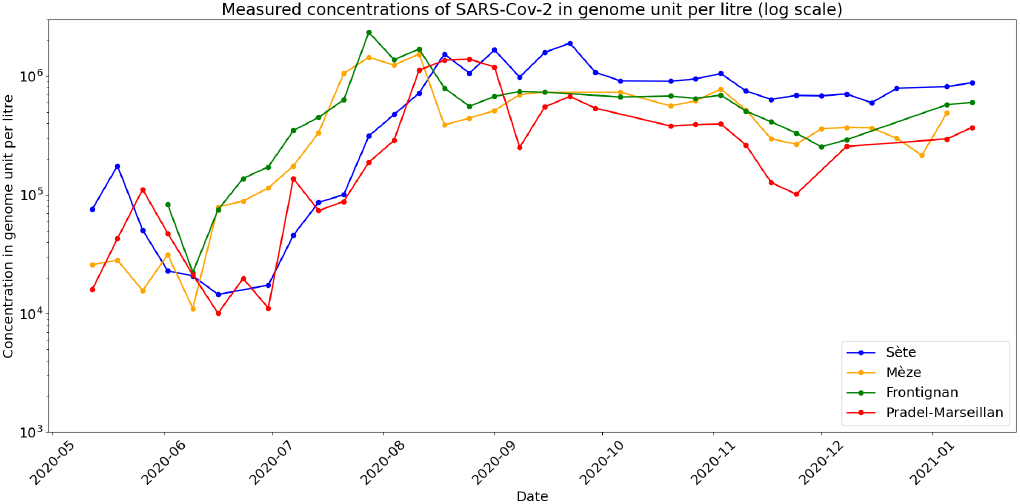
Concentrations of SARS-Cov-2 (genome units per litre in logarithmic scale) from four WWTPs in Thau lagoon, measured weekly with dPCR technology from May 12^*th*^ 2020 to January 12^*th*^, 2021. Note that there are some missing points.

1. An exponential phase starts simultaneously in Mèze and Frontignan WWTPs in early June.
2. The genome units concentration curves in these two places reach, again simultaneously, a plateau. It has stayed essentially stable or slightly decreasing since then.
3. The evolution at Sète and Pradel-Marseillan remarkably followed the previous two zones in a parallel way, with a two weeks lag. The measurements at Sète and Pradel-Marseillan continued to grow linearly (recall that this is in log scale, thus exponentially in linear scale), while the Mèze and Frontignan figures have stabilised ; then, after two weeks, they too stabilised at a plateau with roughly the same value as for the other two towns.
4. The measurements seem to show a tendency to increase over the very last period.

### 4 The epidemiology model with heterogeneity and natural variability of population behaviour

The appearance of such plateaus and shoulders need not be explained either by psychological reactions or by public health policy effects. Indeed, the regulations were roughly constant during the measurement campaign and awareness or fatigue effects do not seem to have altered the dynamics over this long period of time. Witness to this is the fact that two groups of towns saw the same evolution, but two weeks apart one from the other. To understand this phenomena we propose a new model.

Given the complexity and multiplicity of behavioural factors favouring the spread of the epidemic, we assume that the transmission rate involves a normalised variable *a* ∈ (0, 1) that defines an aggregated indicator of risky behaviour within the susceptible population. Thus, we represent the *heterogeneity* of individual behaviours with this variable. We take *a* as an implicit parameter that we do not seek to calculate. The classical SIR model is macroscopic and the type of model we discuss here can be viewed as intermediate between macroscopic and microscopic.

The initial distribution of susceptible individuals *S*_0_(*a*) in the framework of a SIR-type compartmental description of the epidemic can be reasonably taken as a decreasing function of *a*. We take the infection transmission rate *a ↦ β*(*a*) to be an increasing function of *a*. In the Supplementary Information (SI) Appendix, the reader will find a more general version of this model involving a probability kernel of transition from one state to another. The model here can be derived as a limiting case of that more general version.

Likewise, the behaviour of individuals usually changes from one day to another (21). Many factors are at work in this *variability*: social imitation, public health campaigns, opportunities, outings, the normal variations of activity (e.g. work from home certain days and use of public transportation and work in office on others) etc. Therefore, the second key feature of our model is *variability* of such behaviours: variations of the population density for a given *a* do not only come from individuals becoming infected and leaving that compartment but also results from individuals moving from one state *a* to another (21). In the simplest version of the model, variability is introduced as a diffusion term in the dynamics of susceptible individuals.

#### The model

We denote by *S*(*t, a*) the density of individuals at time *t* associated with risk parameter *a*, by *I*(*t*) the total number of infected, and by *R*(*t*) the number of removed individuals. We are then led to the following system:

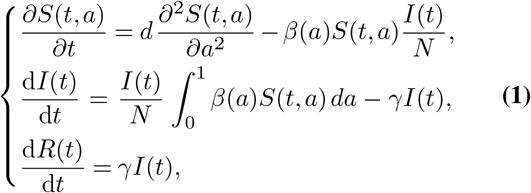

where *γ*^−1^ denotes the inverse of typical duration (in days) of the disease and *d* a positive diffusion coefficient. System (1) is supplemented with initial conditions

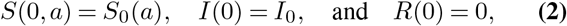

and with zero flux condition in *a* at *a* = 0, 1. In the Method section below, we discuss the relation of this model with other current works.

#### A more general model

In a more general version of our model, we can consider the population of infected as also structured by the parameter *a*. The equations are as before but now we keep track of the class *a* in the infected population. The mechanism here is that a susceptible individual from class *a* can be infected by infectious from any class *I*(*t, b*) but then gives rise to an individual *I*(*t, a*) of the same parent class. We also assume that there is a diffusion of the infected behaviours. We denote by 𝔅(*a, b*) the transmission rate of *S*(*t, a*) by *I*(*t, b*). For simplicity and because it is natural, we will assume that it is of the form

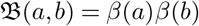

where *β* is as before. For full generality, we can also envision multi-dimensional parameters *a* ∈ ℝ^*d*^, with *a*_*i*_ ∈ (0, 1). We are then led to the system:

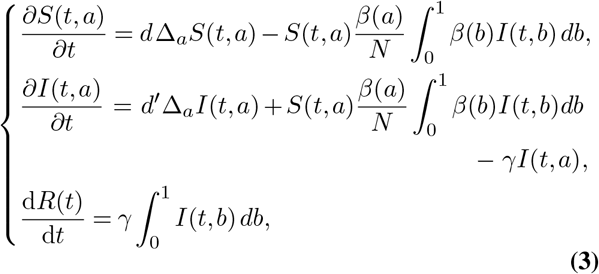

In the SI we write further, more general, forms of this model, with 𝔅(*a, b*) and more general diffusion of behaviours, that can include jumps or non-local variations. The type of models we discuss here may also shed light on the initial phase of the epidemic. We plan to investigate these questions in future work.

### 5 Patterns generated by the model

In the next section, we will discuss how the model fits the data observed in the Thau lagoon measurements. But before that, we start by showing that the above model Eq. (1) can generate the different patterns we mentioned. For this we rely on numerical simulations without fitting real data. And indeed we obtain plateaus, shoulders, and oscillations. The latter can be interpreted as epidemic rebounds.

The key parameter here is the diffusion coefficient *d*, which controls the amplitude of behavioural variability (see Figure 2). Large values of *d* rapidly yield homogenised behaviours, leading to classical SIR-like dynamics of infectious individuals. For very small values of *d*, the system also has a simple dynamics, in the sense that *I*(*t*) has a unique maximum, and therefore has no rebounds. We derive this in the limit *d* = 0 for which we show in the SI that there are neither plateaus nor rebounds.

**Fig. 2.**
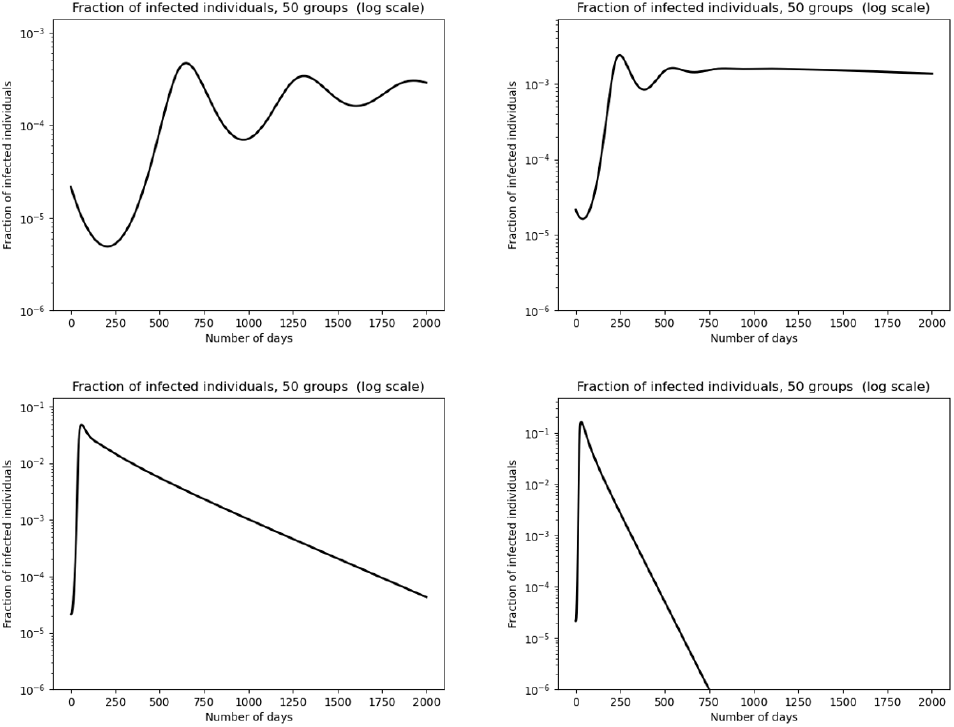
Model behaviour depending on diffusion parameter values: infected rate dynamics in logarithmic scale. From left to right and then top to bottom, graphs are associated with *d* = 10^−5^, *d* = 5 · 10^−5^, *d* = 10^−3^ and *d* = 5 · 10^−3^ (in *day*^−1^ unit).

For some intermediate range of the parameter *d*, plateaus may appear after an exponential growth, like in the initial phase of the SIR model. A small amplitude oscillation, called “shoulder”, precedes a temporary stabilisation on a plateau, followed by a large time convergence to zero of infectious population. We also show that for small enough *d*, time oscillations of the infectious population curve, i.e. epidemic rebounds, may be generated by Model Eq. (1). Such oscillations also appear after a plateau, in a similar way to what one can see in observations.

Simulations in Figure 2 illustrate the various patterns obtained on the dynamics of infected population as a function of the diffusion parameter. For small enough *d*, in the top left graph of Figure 2, one can see oscillations of the fraction of infectious individuals. These oscillations cannot be achieved in the classical SIR model. In fact, the two lower graphs of that figure, for somewhat larger values of *d*, exhibit the SIR model outcomes. Indeed, for sufficiently large *d*, the system becomes rapidly homogeneous (i.e. constant with respect to *a*). Yet, such oscillations are standard in the dynamics of actual epidemics, like the current Covid-19 pandemic. The intermediate value of *d*, represented in the upper right corner of Figure 2 shows the typical onset of a plateau at a rather high value of *I*. Note that this plateau is preceded by a first small dip and then a characteristic “shoulder-like” oscillation.

Secondary epidemic peaks are of lower amplitude than the first one, as shown in the top graphs of Figure 2. This empirical observation leads us to conjecture that, at least in many cases, it is a general property of this model (with *β* independent of time). This property would then reflect a kind of dissipative nature of Model (1). It is natural to surmise that a change of behaviours in time may generate oscillations with higher secondary peaks. Such changes result for instance from lifting social distancing measures or from fatigue effects in the population.

We illustrate this with numerical simulations in Figure 3. We assume a collective time modulation of the *β*(*a*) transmission profile. That is, we replace *β*(*a*) by *β*(*a*)*φ*(*t*) for some time dependent function *φ*, the other parameters are the same as in the simulations shown in Figure 2. We look at the effect of a “lockdown exit” type effect. Then, *φ*(*t*) takes two constant values, 1 from *t* = 0 to *t* = 1000 and 1.2 after *t* = 1100. In between, that is, for *t* ∈ (1000, 1100), *φ*(*t*) changes linearly from the value 1 to 1.2.

**Fig. 3.**
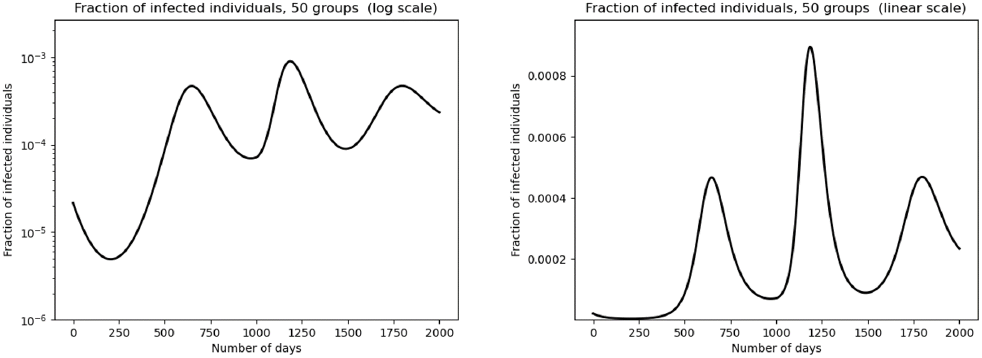
Multiple epidemic rebounds: susceptible individuals is divided into 50 discrete groups in the case where relaxation of social distancing measures starts on Day *t* = 1000 and end up on Day *t* = 1100. The fraction of infected individuals in the population is represented in the left graph in logarithmic scale and in linear scale in the right graph.

One can clearly see a secondary peak with higher amplitude than the first one. Note that after this peak, a third one occurs, with a lower amplitude than the second one. This third peak happens in the regime when *β* is again constant in time.

#### The effect of variants

Another important factor that yields secondary peaks with higher amplitudes is the appearance of variants. Consider the situation with two variants. We denote by *I*_1_(*t*) and *I*_2_(*t*) the corresponding infected individuals. The first variant, which we call the historical strain, is associated with *β*_1_ and *I*_1_(0) and starts at *t* = 0. The variant strain corresponds to *β*_2_ and *I*_2_ and starts at Day *t* = 1000. In this situation, the system Eq. (1) is extended by the following system:

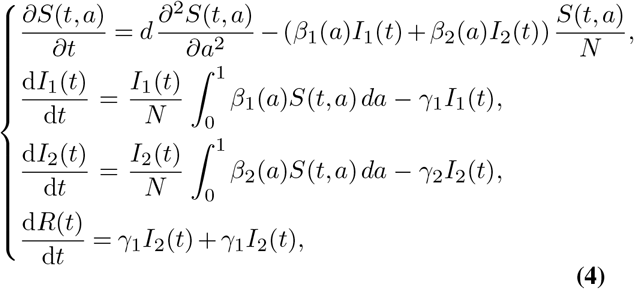

The total infected population is *I*(*t*) = *I*_1_(*t*) + *I*_2_(*t*). Figure 4 shows a simulation of this system. Before the onset of the second variant, i.e. for *t <* 1000, we observe a peak, followed by a small shoulder and a downward tilted plateau. The second variant corresponds to a higher transmission coefficient: namely, we take here 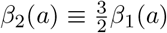. When it appears at time *t* = 1000, initially there is no effect, because the initial number of infectious with variant 2 is very small. Then, there is an exponential growth caused by this second variant gaining strength. The secondary peak is then higher than the first one. A very small shoulder precedes another stabilisation on a downward plateau.

**Fig. 4.**
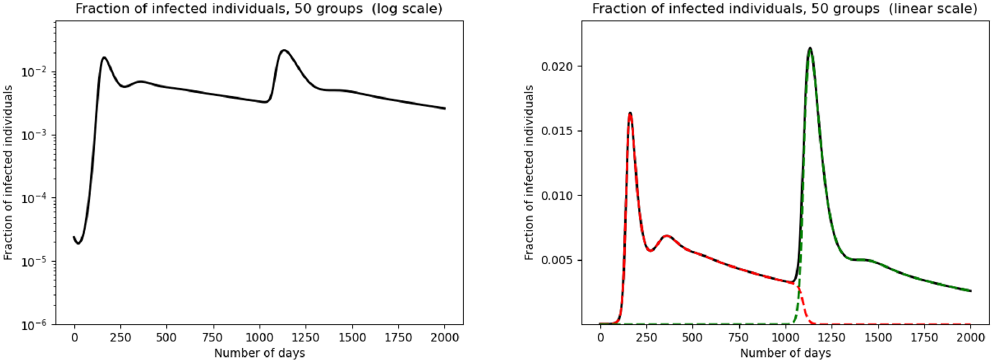
Multiple epidemic rebounds due to a variant virus: susceptible individuals is divided into 50 discrete groups in the case where a new variant appears at Day *t* = 1000. The transmission rate *β*_2_ is taken as *β*_2_(*a*) = 1.5 *β*_1_(*a*), *d* = 0.0002, *γ*_1_ = 0.1 and *γ*_2_ = 0.05. The fraction of infected individuals in the population is represented in the left graph in logarithmic scale. The total infected population is represented in linear scale in the right graph (black curve), variant 1 in red and variant 2 in green.

Figure 4 also shows the dynamics of fractions of infected with each one of the variants. Note that the infectious with variant 1 very rapidly all but disappear at the onset of the second exponential growth phase. One might have expected that the historical strain would be gradually replaced by the new strain, merely tilting further downward the plateau. But that does not happen. Thus, it is remarkable that the historical strain gets nearly wiped out at the very beginning of the second exponential growth.

### 6 Application to the Thau lagoon measurements

Model (1) describes the dynamics of the fraction of infectious in the population, that is *t* ↦ *I*(*t*)*/N*. Therefore, we need to derive this fraction from the wastewater measurements. To this end, we use an “effective proportionality coefficient” between the two quantities. This coefficient itself is derived from the measurements (compare Section “SARS-Cov-2 concentration measurement from wastewater with digital PCR” in the methods part below). Calibration of model (1) also requires fitting the values of *γ*, the profiles *a* ↦ *β*(*a*) and the initial distribution of susceptible individuals in terms of *a*.

We carried this procedure and the resulting fitted curve is displayed in Figure 5. Note that the outcome correctly captures the shoulder and plateau patterns.

**Fig. 5.**
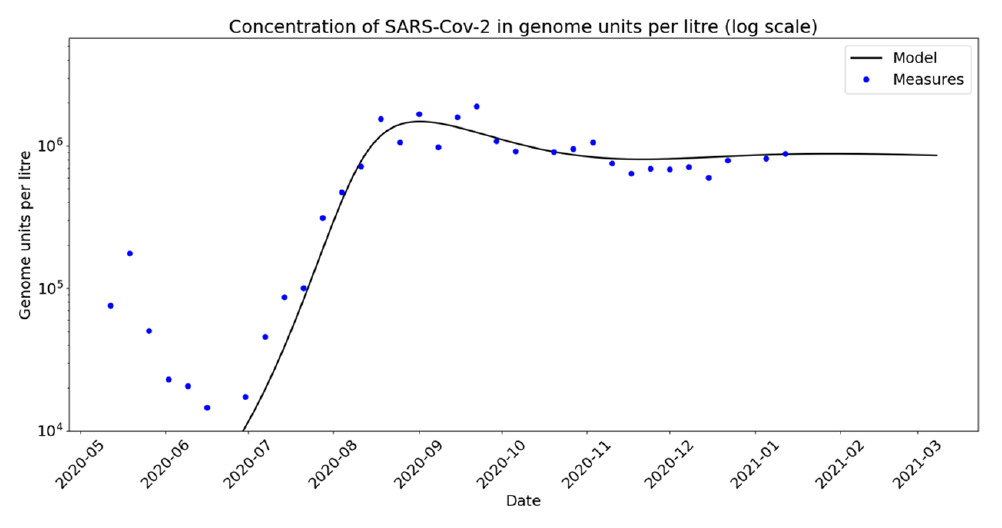
Calibrated model on Sète area: blue dots are measures of SARS-CoV-2 genome units and black curve represents the total infected individuals as an output of the model discretized into *n*_*g*_ = 20 groups in *a*. Initial distribution of susceptible individuals and *β* function are taken as described in supplementary information. Parameters *d* and *γ* are taken as follows: *d* = 2.5·10^−4^ *day*^−1^, and *γ* = 0.1 *day*^−1^.

The underlying dynamics of the rate of susceptible individuals is given in Figure 6 below for *n*_*g*_ = 20 groups. The lower curve illustrates the competition phenomenon between diffusion and sink term due to new infections, depending on the level of risk *a* of each state.

**Fig. 6.**
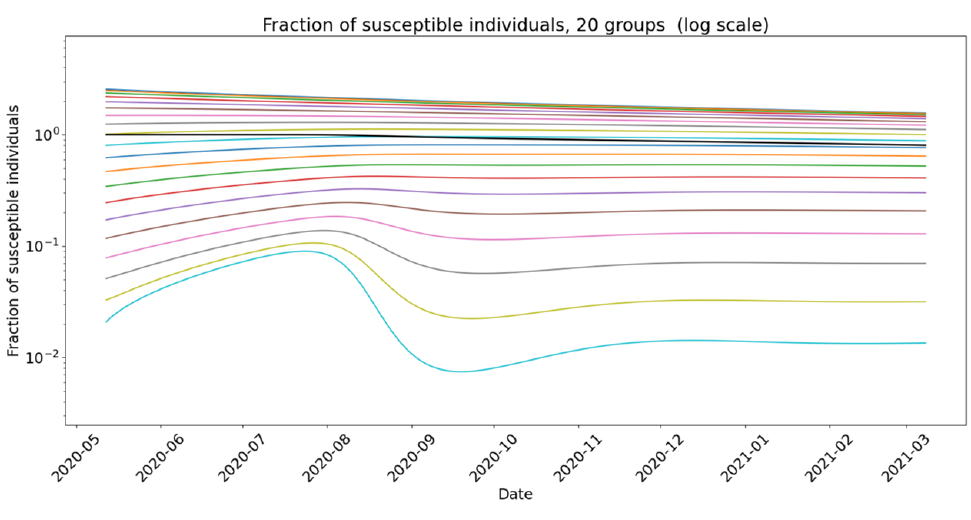
Calibrated model on Sète WWTP: density of susceptible individuals of each group for *n*_*g*_ = 20. The densities of susceptible of each group is represented in colour curves as functions of time. The curves are ordered from top to bottom according to increasing *a*. The resulting average total susceptible population is represented in black. Susceptible individuals of highest *a* trait, which are represented in the bottom light blue curve exhibit a non monotonic behaviour.

### 7 Discussion

We claim that Model (1) explains the formation of plateaus and rebounds in the dynamics of the outbreak through the heterogeneity and variability of population behaviour with respect to epidemiological risk. Figure 6 shows that susceptible individuals with the riskiest behaviour, characterised by the highest *β* transmission coefficient, are rapidly transferred to the infected compartment. Variability of behaviours modelled by diffusion with respect to *a* parameter then refeeds the fringe of riskiest susceptible individuals. Parameter regimes where those two phenomena have the same order of magnitude generate patterns such as plateaus, shoulders and rebounds. We explored above the types of patterns arising when we vary the diffusion coefficient *d*. We note that plateaus occur for an *intermediate range* of values of *d >* 0, which represents the amplitude of variability. Namely, large values of *d* lead to standard SIR-type dynamics, since diffusion quickly homogenises the behaviours. When *d* decreases, plateaus starting with shoulder-like patterns appear. However, for even smaller values of *d*, oscillations arise, which can be interpreted as epidemic rebounds. From numerical experiments, the amplitude of rebounds always seem to be of smaller amplitude than the first epidemic peak. However, higher secondary peaks arise when we significantly modulate in time the transmission rate. This may represent a progressive exit from lockdown or the effect of new and more contagious variants

The dynamics of the Covid-19 outbreak in the cities of Thau lagoon appears almost insensitive to public health regulations. In particular the second lockdown in France from 28 October to 14 December 2020 had hardly any effect. Likewise, the Christmas Holiday season also seemed to have had little influence on the observed plateaus.

The number of hospitalised individuals in France over the last quarter of 2020 is represented in Figure 7. There, the dynamics shows a growing phase followed by a shoulder and a plateau, very similar to the pattern observed in Thau lagoon.

**Fig. 7.**
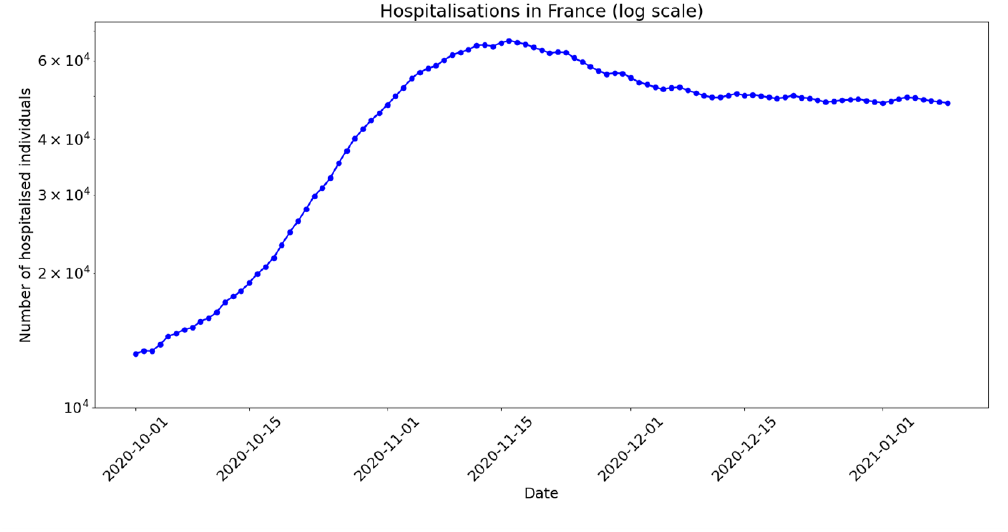
Number of hospitalised individuals in France (log scale, As from October 2020).

Several noteworthy observations came out from the Thau lagoon data. First, we can see two distinct exponential growths in two separate subsets of two towns. The two graphs are parallel with a constant delay of two weeks. The first group reaches a plateau at a certain level of infected and stays thereafter essentially flat, while the second group continues to grow until it reaches about the same value of infected and then becomes essentially flat too. These remarkable observations call for interpretations. Indeed, first, they cannot simply result from public policy measures as these would have affected all these neighbouring towns in a similar fashion. Second, there likely is a spatial diffusion effect that triggered the growth in the second group coming from the first one. However, this diffusion would not explain the fact that the two groups reached the same level of plateaus.

It is worth noting that these observations are inconsistent with the classical SIR model: in this model, once an epidemic reaches a peak, it then decreases steadily by another exponential factor. In other words, a plateau requires an effective ℛ_*t*_ number (22, 23) of approximately one: ℛ_*t*_ ∼1. However this would be hard to sustain over such a long period of time as observed because the susceptible population is depleted. It also appears difficult to explain that this occurs exactly at the same plateau level for two distinct populations.

#### Conclusions

In this paper we reported on precise concentration measurements of SARS-CoV-2 in wastewater upstream of four WWTPs around the Thau lagoon in South of France during the nine months period from May 2020 to January 2021. These observations exhibit plateaus and shoulder patterns. Such characteristics, along with rebounds, are widely observed in epidemics. We provided here an explanation by considering that the population is heterogeneous in the level of risk in the individual behaviours, which random changes from day to day. Indeed, we show here that the combination of heterogeneity and variability leads to a constant replenishment of the population of susceptible individuals with higher risks, thus feeding as it were the epidemic. This mechanism explains the formation of plateaus in the dynamics of the epidemic and also accounts for oscillations and rebounds.

To substantiate this claim, we proposed a mathematical model for epidemics that explicitly involves heterogeneity and variability. The model takes the form of an SIR system with diffusion of behaviours. To get a model as parsimonious as possible (24), we only assumed the diffusion of risks among susceptible and that a single continuous and one-dimensional risk variable *a* characterises the behaviour type. Numerical simulations of this system indeed show exactly this type of shape that we observed, with shoulders preceding long plateaus, and rebounds. Without any change in the behaviours, the secondary peaks, although they can be important, are of a lesser amplitude than the first peaks. Here we show further that a one time change in the transmission coefficients may generate rebounds with higher amplitudes. Such changes typically occur when social distancing measures are lifted.

We also explored the effect of introducing a variant strain of the virus, with a slightly higher transmission rate. We also achieved higher amplitude rebounds in this framework. The analysis of the presence of the two strains shows that the historical strain is replaced quite abruptly by the more transmissible one, earlier than one would guess from its intrinsic evolution.

To further validate this model, we fitted it on the data collected in the Thau lagoon area. Here we did not invoke any external effects such as fatigue or change of public health measures. Indeed, over the concerned period, there were arguably few and minor changes. Furthermore, the two weeks delay between the two curves we observed precludes such effects. Indeed, two towns reached the plateau two weeks earlier than the other two, the latter continued to see an increase, until roughly the same level was reached two weeks later. The calibration of our model on the Thau lagoon observations yielded a very good agreement with the data.

#### Perspectives and extensions of the model

There are several directions to extend our work. First, one can consider more general forms of the model as the one introduced in Eq. (3).

Then, one might envision a more precise approach to the parameter *a* beyond the notion of unit of contact. One can take into account e.g., duration and circumstances of contacts. It appears natural to use “risky sociability” in computing the transmission rate. One may also consider multi-dimensional versions of the parameter. Understanding quantitatively the change of behaviour as a function of this variable *a* may require an interdisciplinary avenue of research.

Likewise, it is quite natural to assume that the variability of behaviour also depends on *a* rather than being uniformly distributed. We discuss such an extension in the SI where we show that it leads to an equation with drift terms. We leave developments in this direction to further work.

Another important aspect that transpires in the Thau lagoon data, is the spatial spreading that takes place in the epidemic. In a recent paper, the first author of this study and collaborators (25) have proposed a model at the country level with a quantitative approach to spatial diffusion in France. The study of diffusion at a smaller scale that we might call mesoscopic and its inclusion in the framework we propose here are promising perspectives.

## Methods

### The epidemiological model with heterogeneity and variability

The model we develop here extends the classical SIR compartmental approach by taking into account heterogeneity and variability of behaviours on the susceptible population. The total population is assumed to be constant equal to *N*. That is, we do not take into account incoming or outgoing populations, nor demographic changes. This is a standard assumption in the Covid-19 studies. Actually, one might want to dispense with it if one is to consider a significant amount of travellers, especially during vacation periods or because there is a lockdown that brings many people to leave large cities.

At time *t*, the population of susceptible is structured by *a* and *t* and is described by its density *S*(*t, a*). We assume that the total number of infected is given by *I*(*t*) and that of the removed by *R*(*t*). We do not distinguish from which population strata the infected individuals come from.

One can think of *a* as a trait parameter roughly describing the level of risk a given susceptible individual is taking with respect to Covid-19. It is normalised so that 0 ≤ *a* ≤ 1, *a* = 0 being associated with very cautious people and *a* = 1 to people with very risky behaviour. This implicit variable represents for instance the number of social contacts per day of an individual, taking into account their length, whether social distancing is observed, if it takes place indoor or outdoor, in a more or less crowded environment, or if individuals wear a mask etc. Thus, it can be seen as a lumped variable that represents a global risk score. In this context, the impact of political restrictions can easily be represented by a modification of the distribution law of the behaviours.

We observe that in this model, we adopt the point of view that the behaviour distribution affects the *risk takers* in the susceptible population rather than the behaviour of infected individuals. Indeed, it appears more natural to consider that the susceptible face an ambient distribution *I*(*t*). For instance, the choice to go to a crowded bar where there *might be* a super-spreader, is reflected in variable *a*. Individuals often vary their behaviour, because for instance of fatigue effects for people who have heeded too strongly social distancing calls or, on the contrary, for people who have been reckless and see other people fall sick and consequently become somewhat more cautious. Thus, the potential reservoir of individuals for a given stratus level *a* is not static and it is more important than would appear at first glance.

Hence, we consider that the variations of *S*(*t, a*) do not only come from individuals becoming infected and leaving that compartment but also results from individuals moving from one stratus to another. Here we assume that this shuffling of behaviours follows Brownian motion. We are then led to System (1) presented in the Result section above. At the end-points of the interval (0, 1) for *a*, we impose homogeneous Neumann (zero flux assumption) boundary conditions:

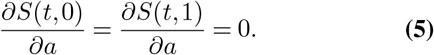

Note that in this system the total population *N* is conserved by the dynamics:

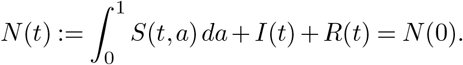

In the SI, we derive this system from more basic considerations and we describe some of its mathematical properties. There, we further discuss more general systems. In particular we consider the framework where the variability itself depends on the trait *a*.

It is enlightening to keep track of the fraction of infected coming from specific strata. To this end, we can introduce a variable *I*(*t, a*) representing the number of infected that came from stratus *a*. It is given by the solution of the equation:

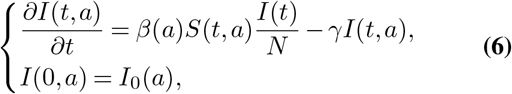

It is straightforward that this is consistent with the definition of the total population of infected, that is:

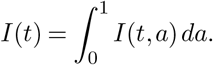

We may include then here the effect that infectious individuals with high *a*’s are more likely to infect other susceptible. This just amounts to replace *I*(*t*) in system Eq. (1)–(6) by

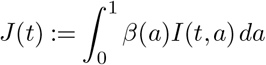

in (1)-(6). We are then led to Model Eq. (3).

### SARS-Cov-2 concentration measurement from wastewater with digital PCR

The teams of the local authorities of the Thau lagoon, *Syndicat Mixte du Bassin de Thau* (or SMBT) collected samples every Tuesday from each of the four WWTPs. Each sample consisted of a compound of 24 hourly samples.

I.A.G.E. (INGENIERIE ET ANALYSE EN GENOME EDITING, 2700 route de Mende 34980 Montferrier-sur-Lez, France) developed a diagnostic method to detect very low concentrations of SARS-CoV-2 in such wastewater samples. This method combines an optimised extraction process with a DNA quantification based on a digital PCR (dPCR) targeting region of the RdRp (IP2/IP4) (20) (This method has been submitted by IAGE to the European Patent Office the 31st of December 2020 under the application number EP20306715.2). The measures produced identical results within three significant digits between the two targets.

In contrast to classical quantitative real-time PCR (qRT-PCR), dPCR allows the *absolute* quantification of low concentration levels of target sequences of nucleic acid molecules from DNA or RNA samples. dPCR outperforms qRT-PCR with respect to accuracy (26) and repeatability of measurements; it is also has a much lower detection threshold (about 20 times lower) (20).

Among recently achieved wastewater measurement campaigns in sewers (27–34), it seems that none of them exploit these measurements in a dynamical model. This is probably due to the uncertainties associated with qRT-PCR measurements and to the difficulty of translating genome unit concentrations into numbers of infected individuals. As outlined in Ahmed et al. (27), the rate of infected individuals within the population served by the instrumented WWTP may be related to the measured genome unit concentration through a proportionality relation:

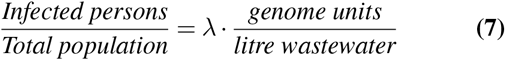

where

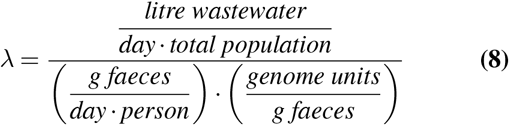

Still, the individual variability of each parameter of Equation (8) is very high, as pointed out in several works (27, 35, 36) (see also references therein). Moreover, transport of wastewater from the emission point to the WWTP involves additional significant phenomena identified in Hart et al. (37): virus degradation over time, usually modelled by exponential decay law where the half life depends on temperature. Therefore, a bottom-up approach estimating each component of Equation (8) from literature is not realistic because of the huge variability and uncertainties of these factors.

Instead, we develop here an original approach to estimate an *effective* value parameter *λ* by viewing *λ* as one of the parameters in the optimisation process in order to fit the model to the data. The reason why we are thus able to determine *λ* lies in the richness of the data set combined with the complex dynamics allowed by the nonlinear model (1). Indeed, the capture of the dynamics of concentrations by the model, including the exponential growth phase, followed by the formation of shoulder-like and plateau patterns, adequately constrains the estimation of the value of this effective parameter *λ*. We computed this parameter for the WWTP of Sete by a least square minimisation process over the time interval from 9 June 2020 to 12 January 2021 covering the preceding three phases. Indeed, since we are interested in these phases, for the sake of clarity we chose to ignore the observations prior to this period. This procedure led us to the choice of *λ*^−1^ = 111, 230, 001 (in genome unit per litre), a figure that seems of the correct order of magnitude.

By this data-driven optimisation procedure, we thus relate the rate of infected people to the measured concentration of SARS-Cov-2. We plan to further investigate and extend this approach in future work.

Such virus concentration time series, essentially proportional to the fraction of people infected by Covid-19, provides an accurate quantitative method to monitor the epidemic. Furthermore, as well known (29, 32, 34), the appearance of the virus in wastewater precedes the observations of the disease and therefore yields a remarkable early warning system, ahead of hospital counts. Moreover, it reflects all infectious people regardless of whether they are symptomatic or asymptomatic.

## Data Availability

All data and codes will be made available upon publication in a journal.

## Acknowledgements

The authors are grateful to the municipalities surrounding the Thau lagoon and to the company “Ingénierie et Analyses en Génétique Environnementale” (I.A.G.E.) for giving them access to the measurements they carried out and which are quoted in this publication.

The authors are also thankful to Jianhong Wu, Marc Barthélemy, Odo Diekmann and Jose Scheinkman for useful discussions about this work.

## Supplementary Information

### The classical *SIR* model with time dependent transmission rate

The classical SIR compartmental model (38) uses a dynamics governed by mass-action laws. It assumes that individuals are homogeneously mixed and that every individual is equally likely to interact with every other individual. It reads:

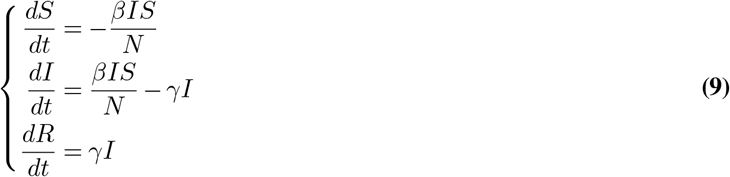

#### Single location growth and behavioural changes

Changes of behaviour driven by phenomena such as awareness of fatalities or fatigue with respect to mobility restrictions can be modelled by a time modulation of the infection rate *β*.

To start with, we analyse this problem on synthetic data. Namely, we consider an exact dynamics given by the SIR model with parameter changes on two given dates. We now take part of the resulting points as given observations. The goal is to carry an optimisation procedure where we try to identify the dates and magnitudes of these changes i.e. the different values of *β* that come into play. Later on we will consider the Thau lagoon data.

Figure 8 represents a piece-wise constant modulation in time with reduction of social interactions down to 70% starting Day *t* = 25 followed by an increase to 50% on Day *t* = 50.

**Fig. 8.**
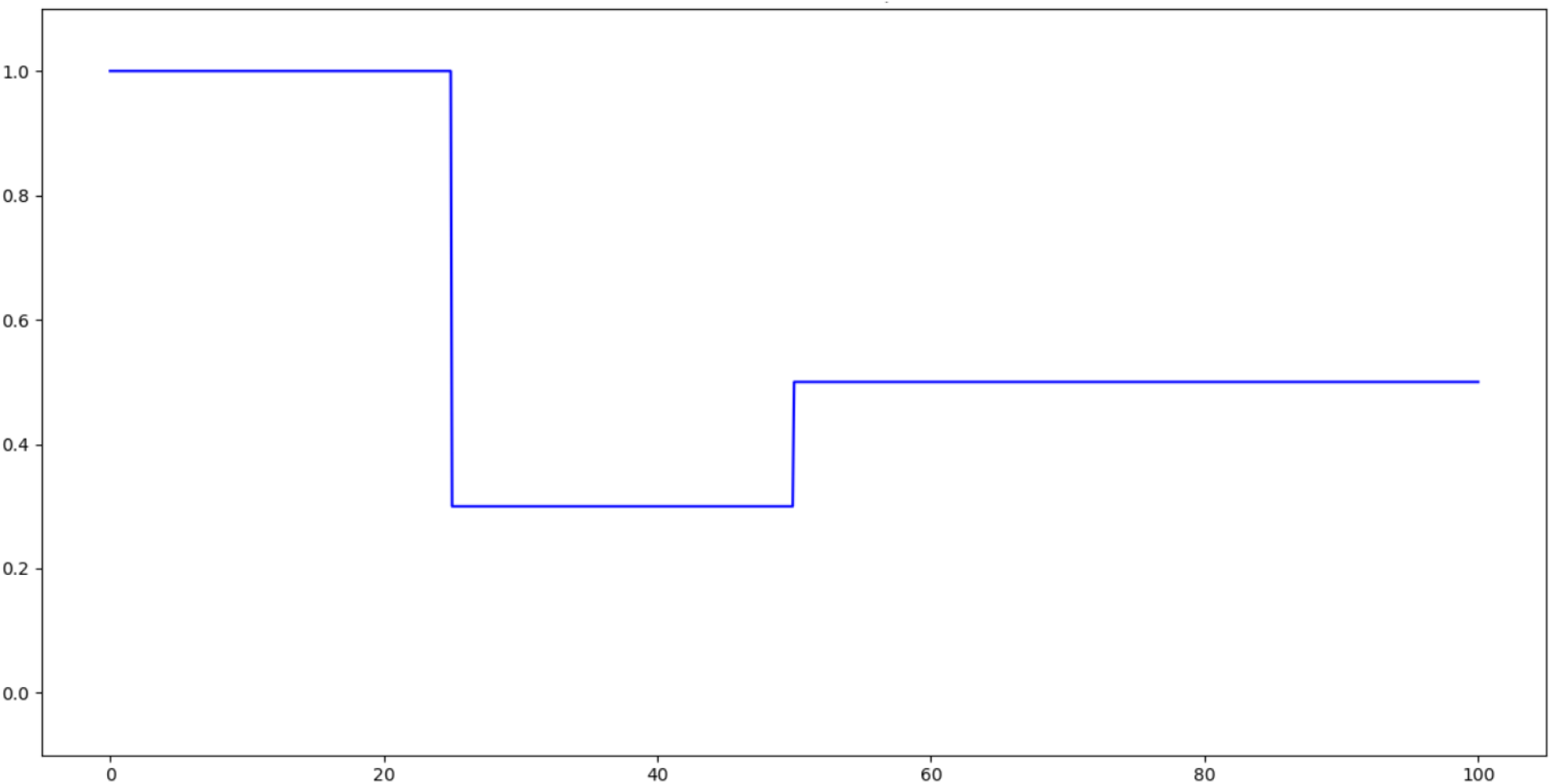
Time modulation function associated with collective behaviour modulation as a function of time (day numbers)

The effect on such modulation leads to the interruption of the growth phase followed by a lower decrease for later times.

We developed a toolbox based upon PYGMO (39) particle swarm optimisation algorithms in order to solve the type of inverse problem we encounter here. We consider a time sampled version of infection rates in the total population. We allow a given number of transition times associated with behaviour changes, in between which the coefficient *β* is constant. The problem then is to determine in an optimal way the SIR model parameters, initial conditions, the transition times and the values of the *β*’s in between. Using a least square objective function, we obtained a satisfactory convergence towards the dynamics of the initial model and associated modulation function.

We developed a similar approach with noise added to sampled synthetic data, with satisfactory convergence both in terms of dynamics of infection rate (Figure 12) and modulation functions (levels and time changes in Figure 13).

**Fig. 9.**
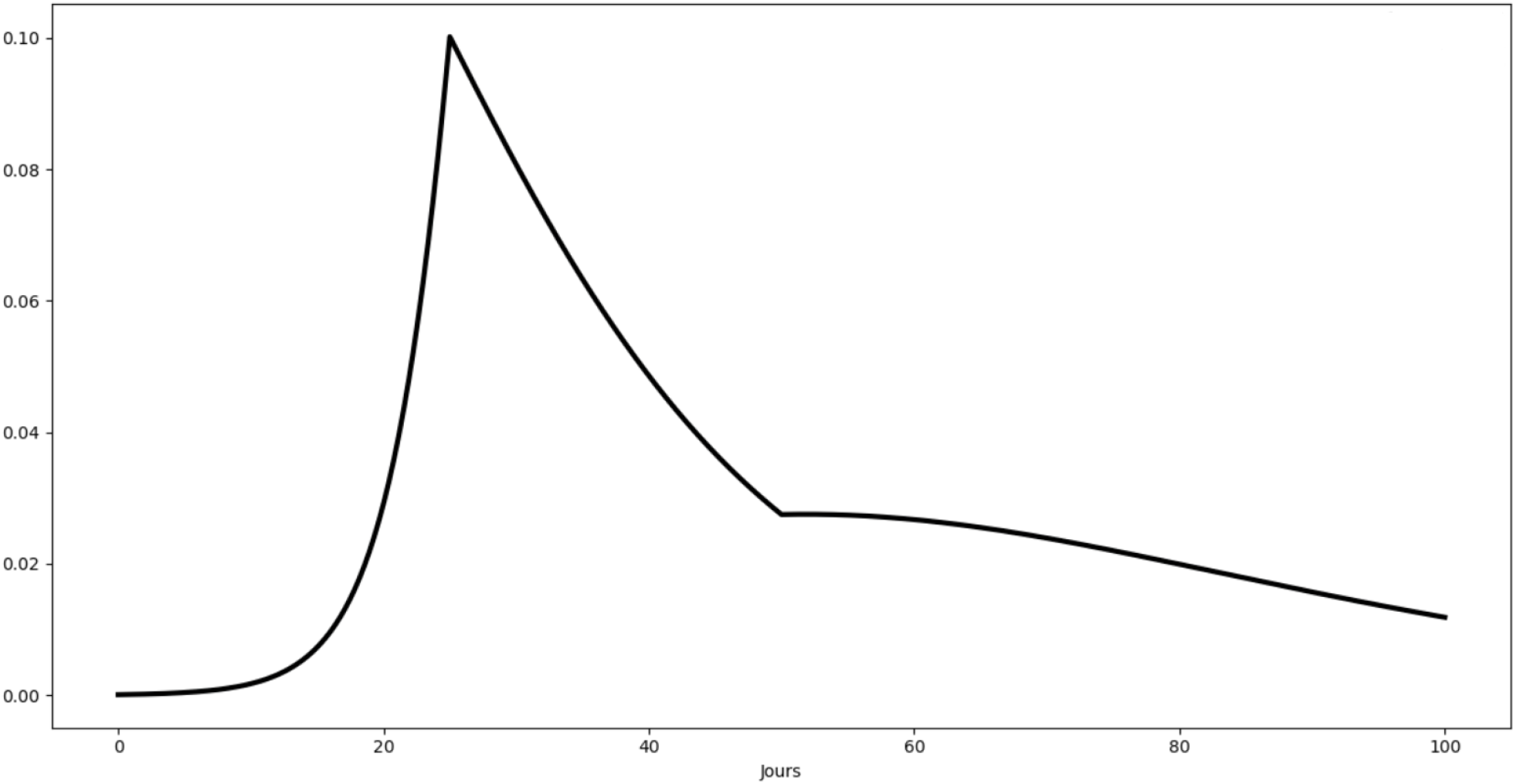
Rate of infected individuals as a function of days

**Fig. 10.**
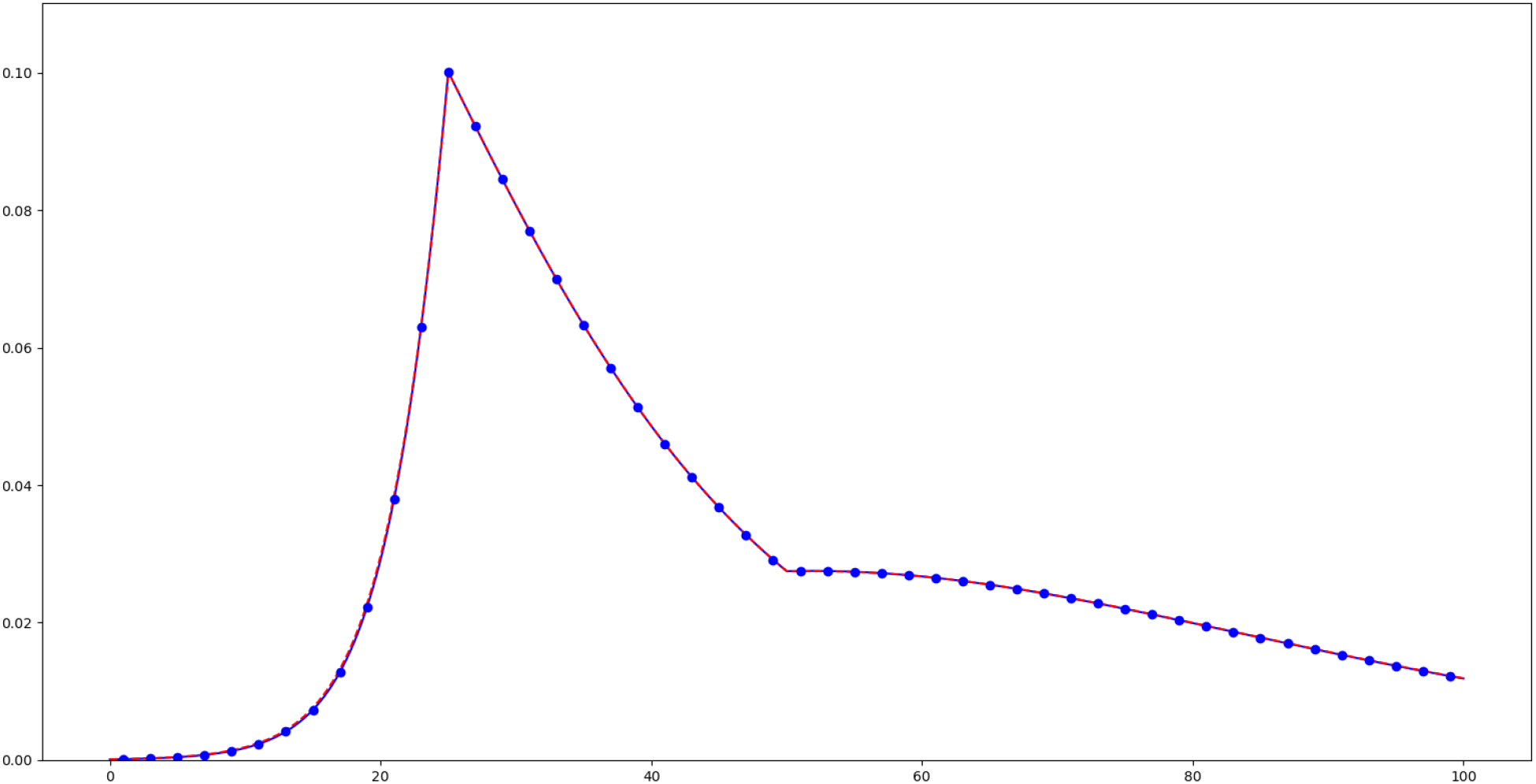
Rate of infected individuals as a function of days as a result of the inverse problem: blue dots correspond to daily sampling of synthetic data, red line to the resulting SIR model.

**Fig. 11.**
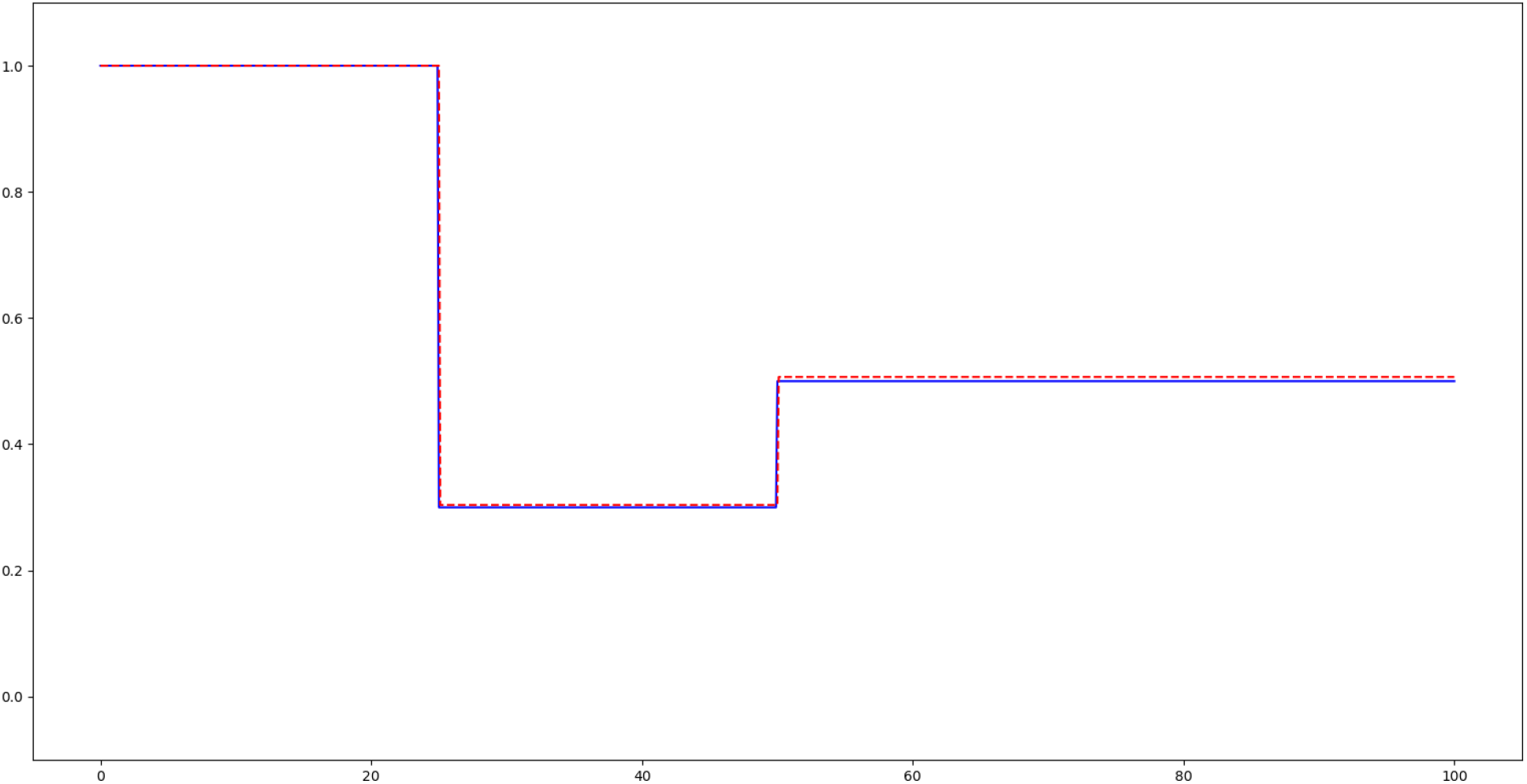
Modulation of collective behaviour as a result of the inverse problem

**Fig. 12.**
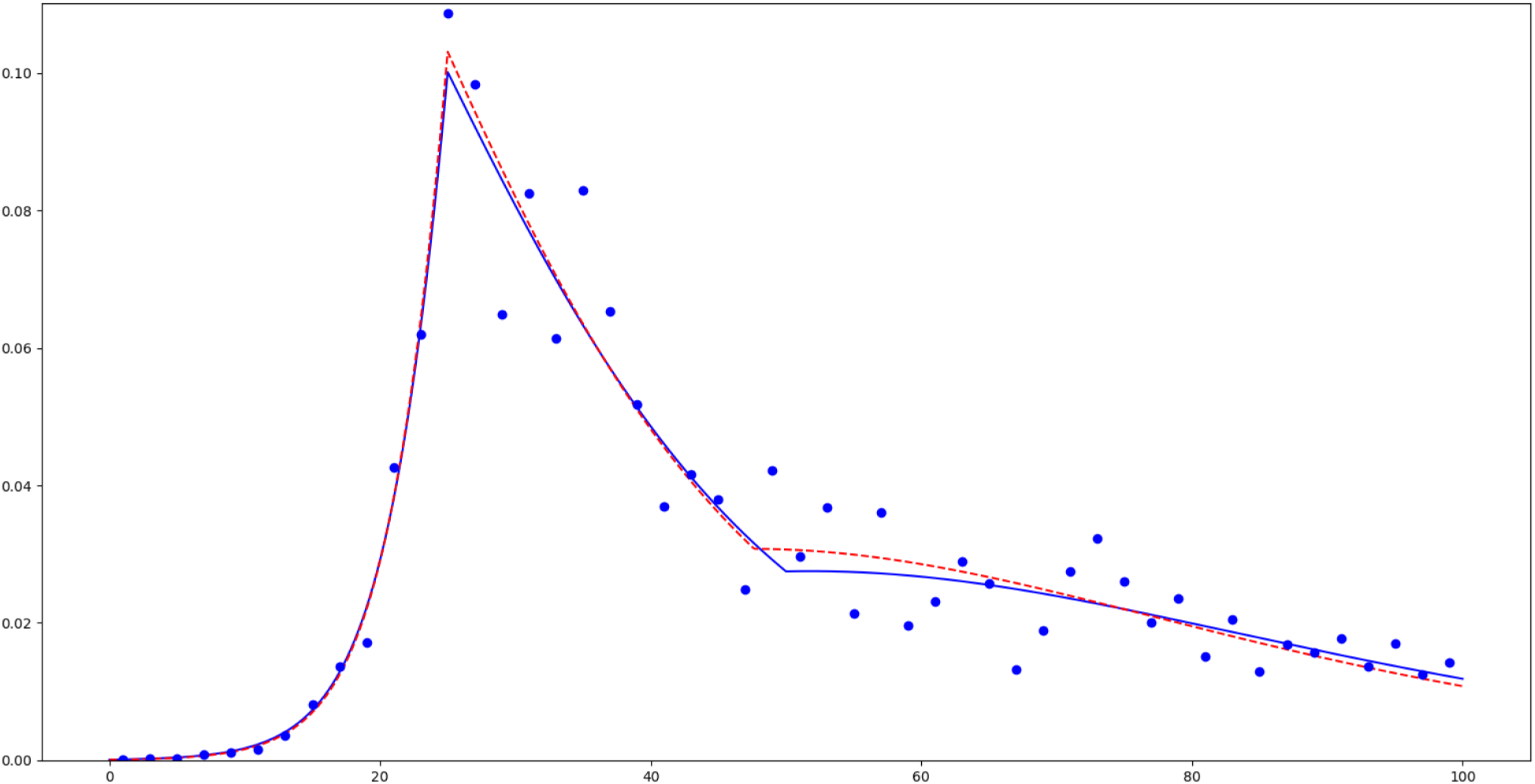
Inverse problem with noisy synthetic data: blue dots correspond to daily sampled synthetic data with noise, blue and red curves respectively to the initial synthetic model and to the result of the inverse problem.

**Fig. 13.**
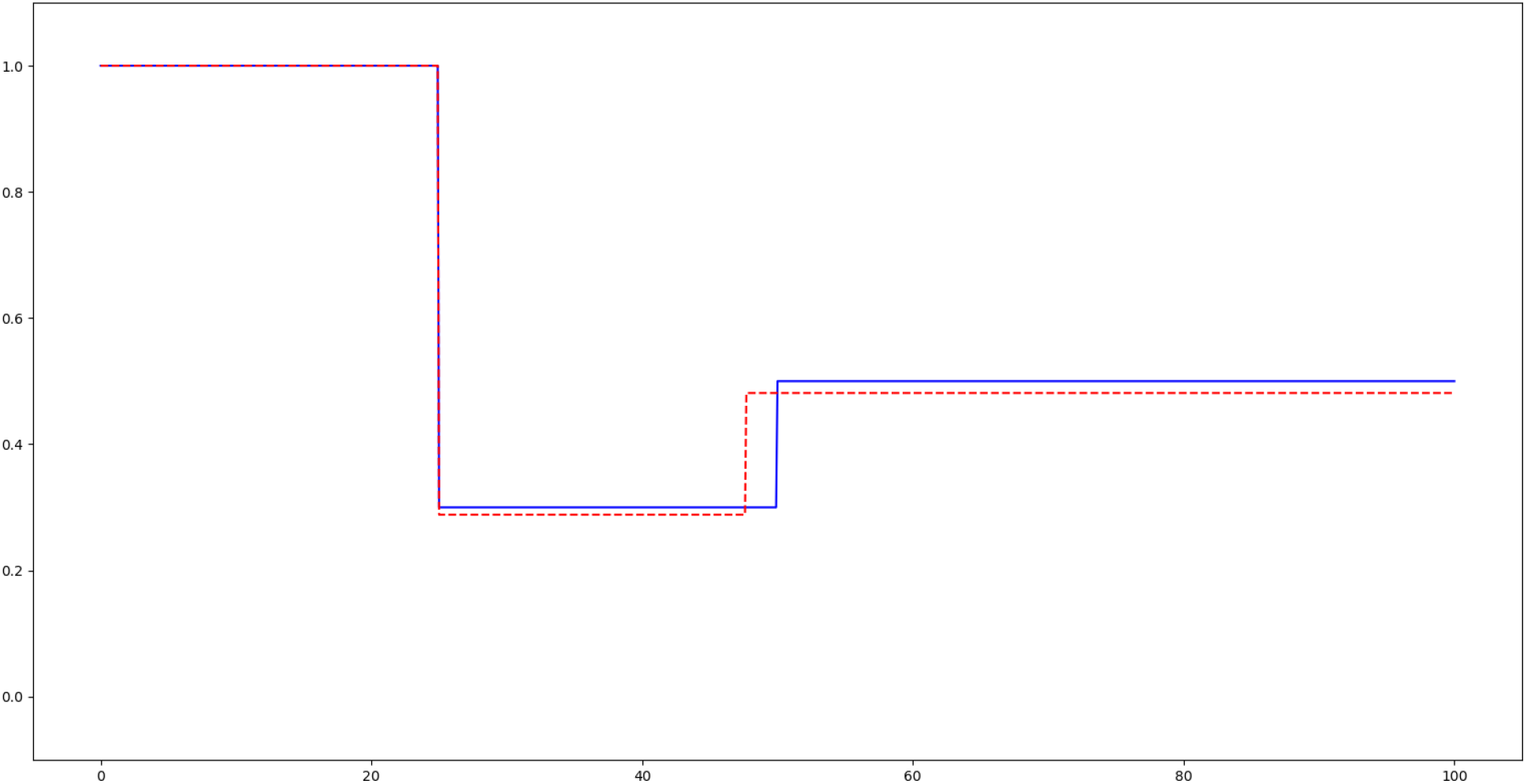
Original modulation function in blue and result of the inverse problem in red.

#### A model with change of behaviour at discrete times in Thau lagoon

We applied the preceding algorithmic approach on real data of the four WWTP measurement campaigns of the Thau lagoon. In order to keep the model parsimonious, we used the same time modulation function of *β* for all the four cities together. The transition dates and levels have to be determined.

The above results show that the optimal capture of the transition from exponential growth to plateau type dynamics occurs on August 10^*th*^, 2020. The resulting piecewise constant modulation function in time is represented in Figure 15 below.

Even though the plateau dynamics seems to be correctly represented in Figure 14, the main shortcoming of this is that it does not really explain the observations of the Thau lagoon data. Indeed, the model does not explain why the four zones reach the same infection rate level associated with the plateau regime and why there is a two weeks delay. Furthermore, it does not yield a shoulder effect, as seen in the data.

**Fig. 14.**
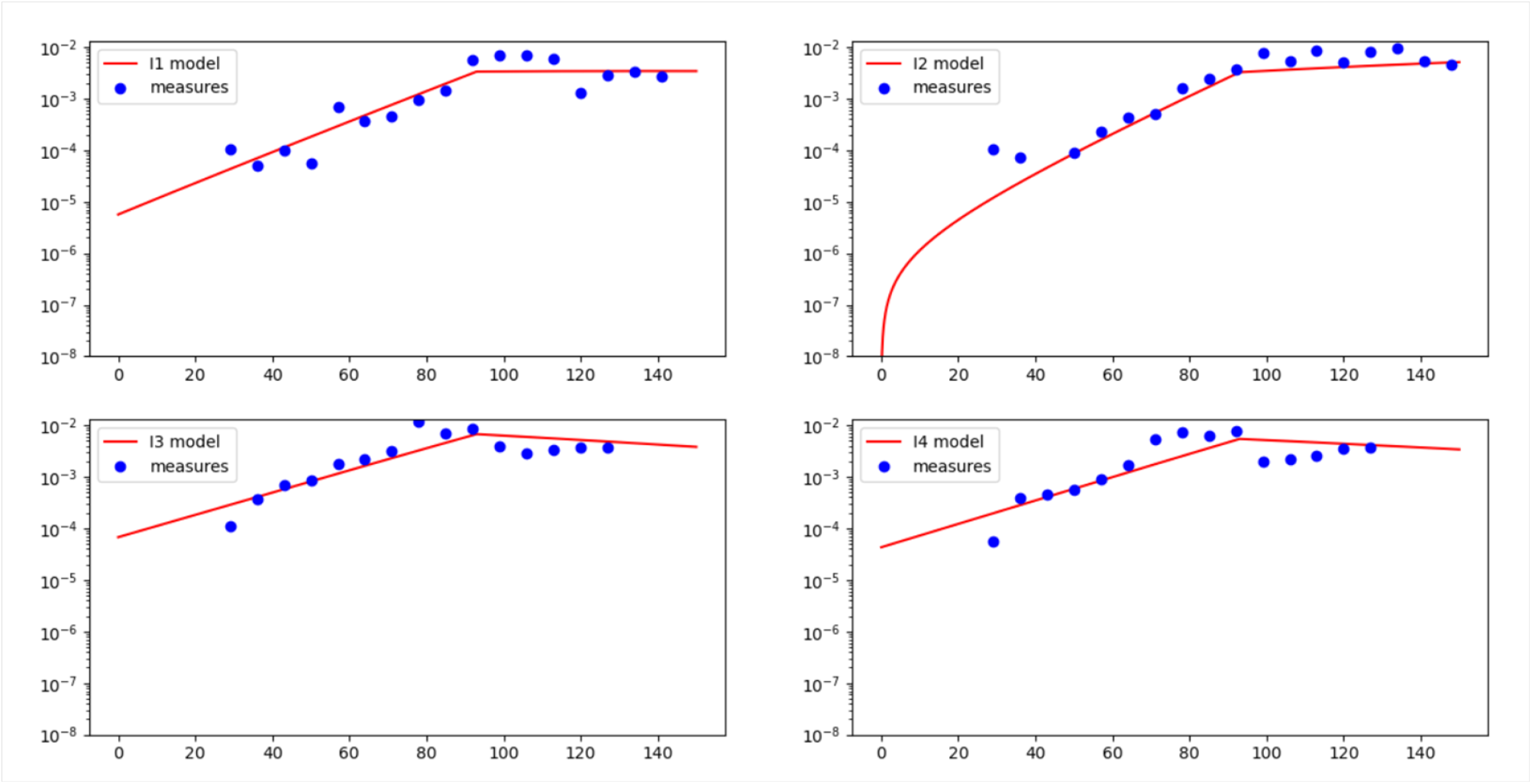
Change of behaviour in logarithmic scale in the Thau lagoon (initial day *t* = 0 being associated with May 12^*th*^, 2020). The red curve represents the model output (rate of infected population) and the blue dots correspond to measurements. Zones *I*_1_, *I*_2_, *I*_3_ and *I*_4_ respectively correspond to Sète, Mèze, Frontignan and Pradel-Marseillan.

**Fig. 15.**
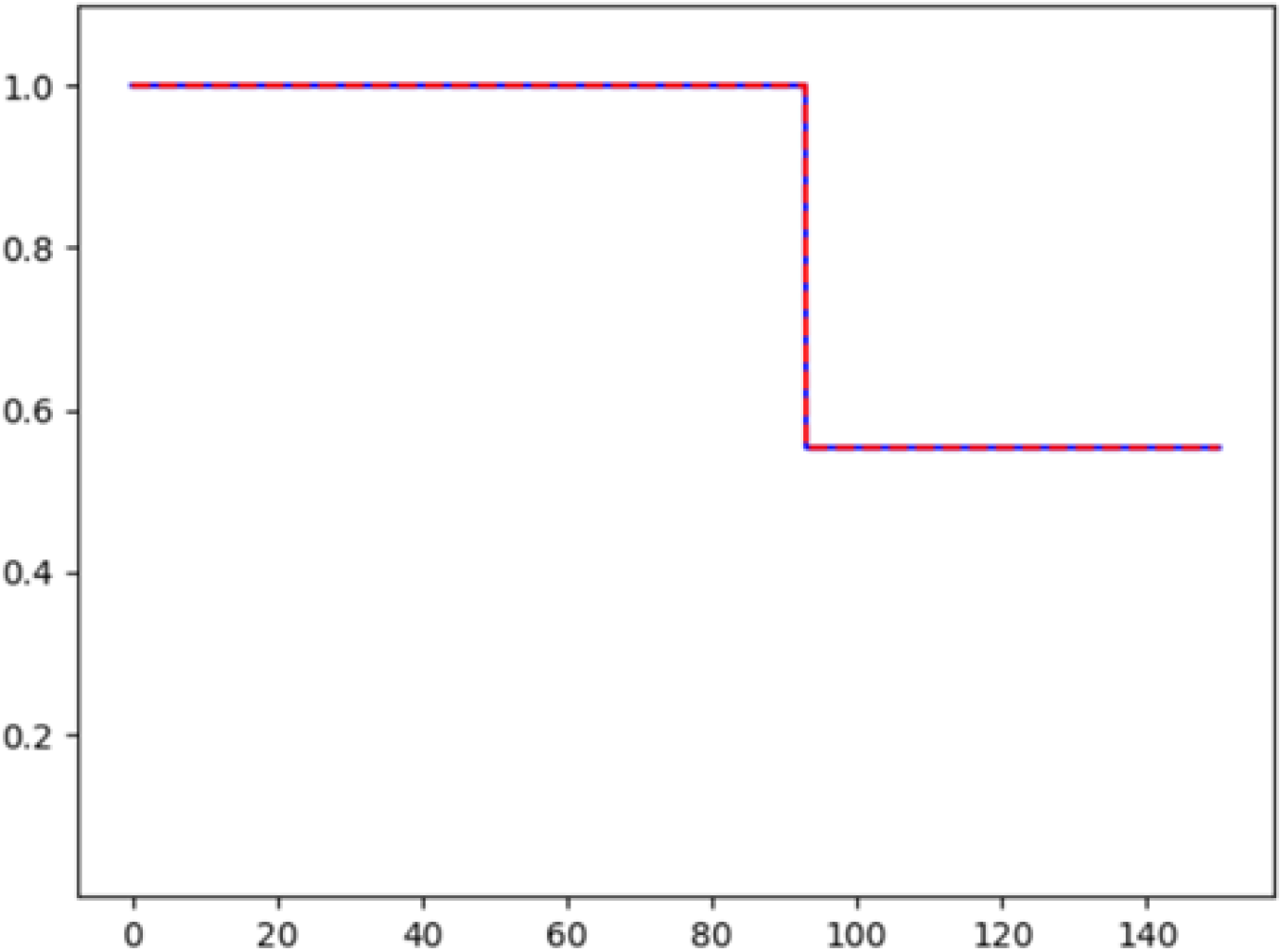
Time modulation of collective behaviour in Thau lagoon as a result of the inverse problem.

One may use spatial diffusion to describe the delay between the zones (Frontignan, Mèze) on the one hand and (Sète, Pradel-Marseillan) on the other hand. But it also fails to explain why there are two parallel curves. We plan to come back to spatial diffusion in future work.

### Derivation of the Model

This section is concerned with the formal derivation of the additional diffusion term in the dynamics of susceptible individuals. As previously emphasised, this diffusion term stems from the combination of the heterogeneity of behaviours and their variability in time. A natural case to consider here is to assume that the shuffling of behaviours happens according to Brownian motion. Namely, individual risk traits move according to the process

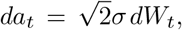

where *σ >* 0 is the possibly *a*-dependent diffusion, and *W*_*t*_ is a Brownian motion in (0, 1) with reflection conditions at the end-points of the interval. By the Fokker-Planck equation, in the absence of epidemic, an initial distribution of population *S*(0, *a*) gives rise to *S*(*t, a*) = 𝔼 [*S*(0, *a*_*t*_)] governed by

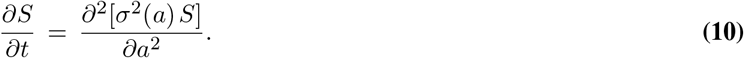

To consider the dynamics of the epidemic, we are thus led to the following more general version:

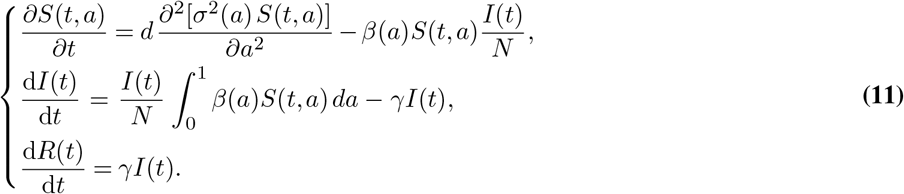

The first equation involves drift and other additional terms. The numerical and theoretical analysis of the case where *σ* depends on a, and the corresponding extensions of the Fokker-Planck equation with drift terms, seem relevant from a modelling viewpoint.

In the simple case where *σ*^2^(*a*) = *d* where *d* is a constant, one ends up with the first equation of our system (see System (12) below).

One could also envision a multi-dimensional variable *a* = (*a*_1_, …, *a*_*p*_) to encompass various behavioural characteristics that could possibly be related to sociological observations. In this case, one would be led to a higher dimensional partial differential equation. where the term 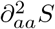 is replaced by the Laplace operator Δ_*a*_*S* as we did in the general model presented in Methods section. Such developments would be useful to improve the model and its applicability.

### Simple properties of Model (12)

For the convenience of the reader, we recall the model introduced in the main paper

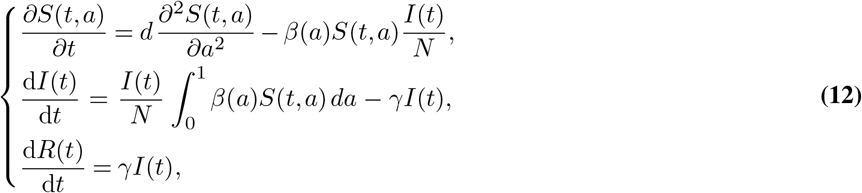

The case *d* = 0 (heterogeneous but without variability) writes:

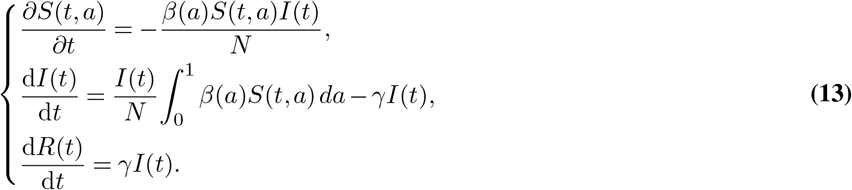

We list below some elementary mathematical properties of the model. First, we note that this model contains the traditional SIR model when the initial profile of susceptible *a* ↦ *S*_0_(*a*) is uniform in *a*. Indeed, it is straightforward to see that then, *S*(*t, a*) also does not depend on *a*.

The dynamics of total infectious individuals is governed by

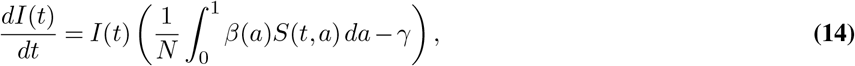

so that the equivalent of the so called “effective R” coefficient in traditional SIR model, namely *βS*(*t*)*/*(*γN*), is replaced by

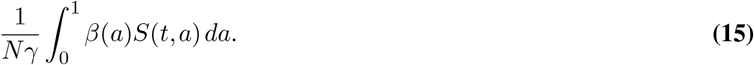

Unlike traditional SIR model, in which the evolution of infectious *t* ↦ *I*(*t*) begins by an exponentially growing phase, has a unique maximum and tends to zero for large time, Model (12) may exhibit more sophisticated behaviours such as rebounds in *I* with multiple local maxima as well as plateaus. This property can be intuitively understood by analysing the evolution of the growth factor in (14):

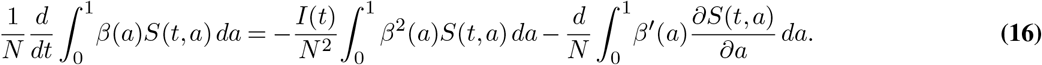

In the absence of diffusion, *d* = 0, this growth factor is non increasing in time. For heterogeneous profiles and non zero diffusion, the first term of the right hand side is always negative whereas the second term may be positive if for instance *a* ↦ *β*(*a*) is increasing and *a* ↦ *S*(*t, a*) is a decreasing function. Below we show that it is always the case under some conditions. Thus, depending on the amplitude of the above two terms, the growth factor may have increasing and decreasing phases. It naturally leads one to consider initial conditions characterised by the property 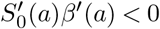 such as

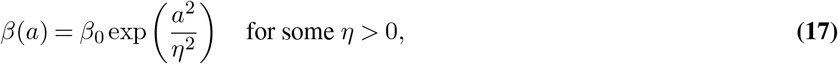

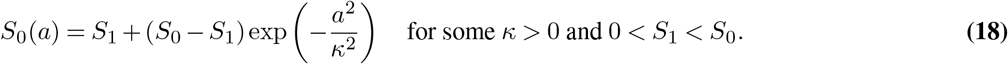

It means that individuals are sorted by increasing infection rate, according to the variable *a*, and that the vast majority of individuals have low infectiousness whereas few individual close to *a* = 1 have extremely risky behaviour. To some extent, social contact surveys (5, 6, 21) justify an initial distribution of susceptible individuals like (18). However, we are not aware of a rigorous justification of the expression (17) of the sharply increasing profile in *β*.

Let us now show that the evolution preserves the property of being decreasing in *a* for the profiles of the susceptible population.

#### Proposition 1.

*If S*_0_ *is a decreasing function of a, then S*(*t*, ·) *is also decreasing in a for all time t*.

To prove this property, we simply note that the derivative *v*(*t, a*) := *∂S*(*t, a*)*/∂a* of *S* with respect to *a* satisfies the following equation

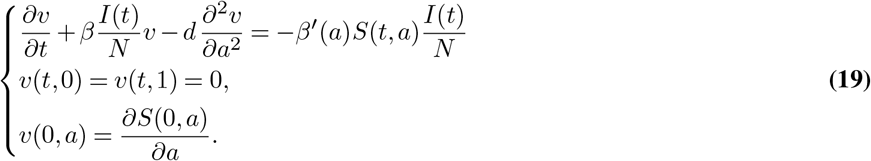

Since the right hand side of the first equation of (19) is negative and the initial condition *v*(0, *a*) ≤ 0, the parabolic maximum principle then shows that *v*(*t, a*) ≤ 0 for all further time.

### Further mathematical properties of Model (12)

Let us first notice that we can describe the dynamics of susceptible individuals in (12) in terms of its probability density *f* (*t, a*) = *S*(*t, a*)*/S*(*t*) where 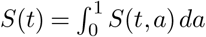:

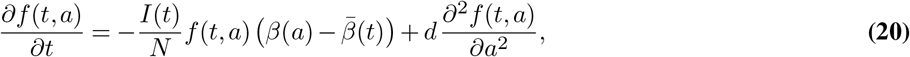

where we denote 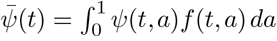. This equation shows the effect of infections on the distribution of susceptible population as a function of *a*. Indeed, it shows that, for instance in the absence of diffusion (*d* = 0), the proportion of the population of high *a* goes down as a result of infection affecting it more in relative terms than the remaining of the population. The presence of diffusion (*d >* 0) mitigates this effect by fuelling as it were the epidemic with individuals who had initial lower risk trait.

The equivalent of (16) for the average transmission rate 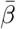 is

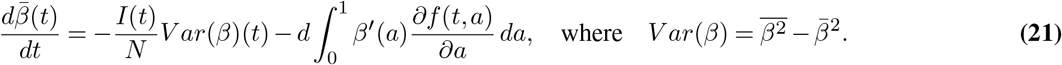

In other words, in the absence of behavioural variability (*d* = 0), the average of the transmission rate decreases under the effect of its variance, the latter being directly linked to behavioural heterogeneity. Thus a higher heterogeneity promotes a faster decay of the average transmission rate. This is a remarkable property of the system in absence of diffusion or with small diffusion. In the presence of diffusion *d >* 0, this effect is in competition with the second term of the right hand side of (21) which may be positive if for instance the profile *a* ↦ *β*(*a*) is increasing while the distribution *f* decreases along variable *a*.

Our next result concerns the effect of heterogeneity on herd immunity. We analyze it in the absence of diffusion.

#### Theorem 1.

*Let* (*t, a*) ↦ (*S*(*t, a*), *I*(*t*), *R*(*t*)) *be the solution of (13) (that is when d* = 0*) with initial conditions* 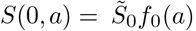, *I*(0) = *Ĩ*_0_ > 0 *and R*(0) = 0 *(for some positive constants* 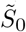 *and Ĩ*_0_*). Then, t* ↦ *S*(*t*, ·) *tends as t* → +∞ *to a limit profile a* ↦ *S*_∞_(*a*) *in such a way that*

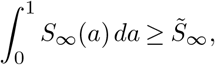

*where* 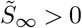 *is the limit value as t goes to* +∞ *of susceptible individuals* 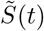 *associated to the homogeneous SIR model (9) with initial conditions* (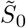, *Ĩ*_0_, 0) *and transmission rate* 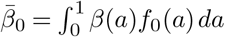.

Thus, this theorem asserts that heterogeneity lowers the herd immunity rate needed to stop an epidemic. Note that Several works study the asymptotic states of similar models without diffusion, in a discrete class framework. We mention in particular Magal et al. (16), Dolbeault and Turinici (14) and Almeida et al. (17).

### Sketch of proof

The existence of the limit profile *a* ↦ *S*_∞_(*a*) follows from the monotonicity of *t* ↦ *S*(*t, a*) for fixed *a* ∈ (0, 1). Since *t* ↦ *I*(*t*) can be easily proved to vanish for large time, the recovered compartment *R*(*t*) also tends to a limit *R*_∞_. Using *R* as a time variable leads to:

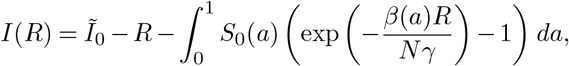

so that

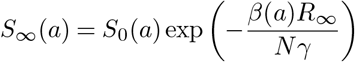

and

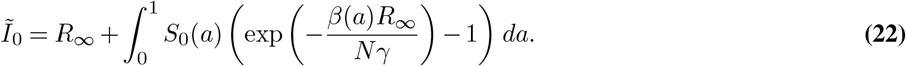

Equation (22) expresses *Ĩ*_0_ as an increasing function *F*_*β*_ of *R*_∞_, which is therefore uniquely defined. Application of Jensen’s convexity inequality then leads to

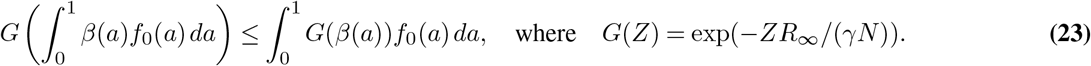

We deduce that 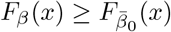 for all *x >* 0, where 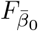 is associated with the homogeneous SIR model (9) with initial conditions 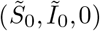 and homogeneous parameter 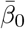, which allows to conclude.

An interesting property of System (13) is the conservation of the following entropy-like quantity

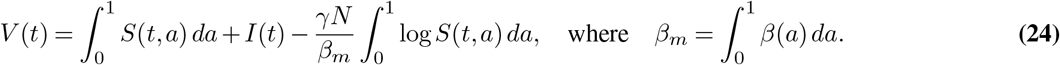

The property *V* (*t*) = *V* (0) can be deduced from (13).

Next, we consider more specifically the role of diffusion *d >* 0. First, we note that in the presence of diffusion, the above entropy *V* ^*d*^ based on solutions (*S*^*d*^(*t, a*), *I*^*d*^(*t*), *R*^*d*^(*t*)) of (12) is non-decreasing in a similar manner as in the framework of viscous perturbations of scalar conservation laws:

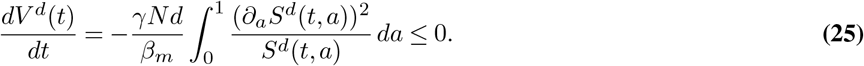

The following result emphasises the differences between the case with diffusion (*d >* 0) and the case without (*d* = 0).

#### Proposition 2.

*Assume d >* 0 *and consider solutions* (*S*^*d*^(*t, a*), *I*^*d*^(*t*), *R*^*d*^(*t*)) *of (12). The following properties hold:*

i. *t* ↦ *S*^*d*^(*t, a*) *is not necessarily decreasing in time for a* ∈ (0, 1) *given*.
ii. *If* 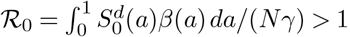, *then there exists t*_0_ *>* 0 *such that t* ↦ *I*^*d*^(*t*) *is increasing for t* ∈ (0, *t*_0_).
iii. *If ℛ*_0_ *<* 1, *t ↦ I*^*d*^(*t*) *is decreasing in time over a finite time interval. Still, there are situations where I*^*d*^ *can be increasing over some later time intervals*.

We will give the proof of this result in a forthcoming work.

We now compare the combined impact of heterogeneity and variability compared with the case of homogeneous populations.

#### Theorem 2.

*For given d >* 0, *let* (*S*^*d*^(*t, a*), *I*^*d*^(*t*), *R*^*d*^(*t*)) *be solution of (12) with initial conditions* 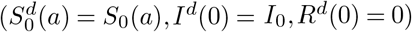. *The following results holds*

i. *S*^*d*^(*t*, ·) *converges as t goes to* +∞ *to a positive constant* 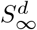 *independent of a*.
ii. *Let* 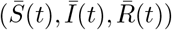 *be solution of the homogeneous SIR model (9) with initial conditions* 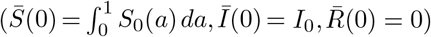 *and transmission rate* 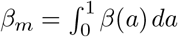. *Then, denoting* 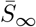 *the large time limit of* 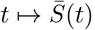, *one has* 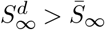.

We already know from Theorem 1 above that heterogeneity is beneficial in terms of herd immunity. From this point of view, the effect of variability is a priori not so clear because the constant shuffling of population keeps fuelling as it were the epidemic. However, part (ii) in the previous result shows that heterogeneity and variability still have a positive effect on herd immunity (that is 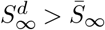).

The convergence of solutions to states that do no depend on *a* is natural because of the role of diffusion. Indeed, the diffusion term takes over once the *I*(*t*) term becomes small and thus the first equation describes a diffusion. But this only happens in the long term and the relevance here is rather at intermediate times.

### Sketch of proof

Assuming that the initial profile *S*_0_ and *β* are regular enough in the *a* variable, integration by parts of the *S* equation of (12) multiplied by *∂*_*t*_*S* leads to:

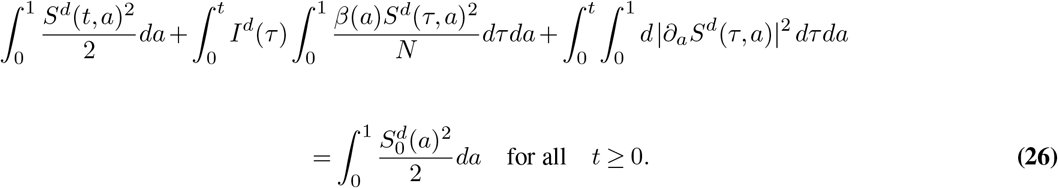

Moreover,

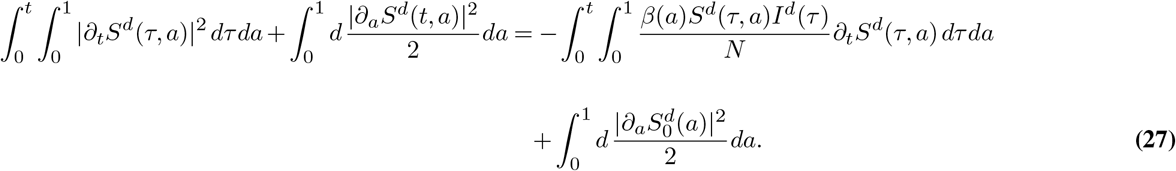

Hence,

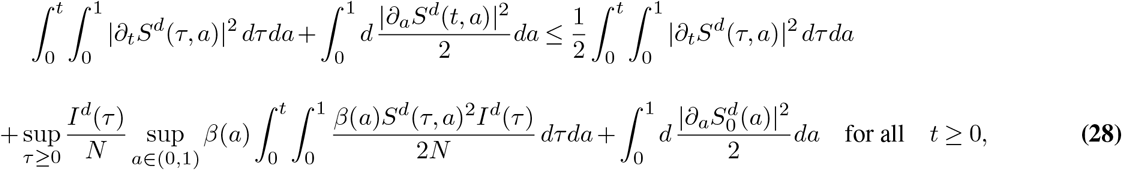

so that we have

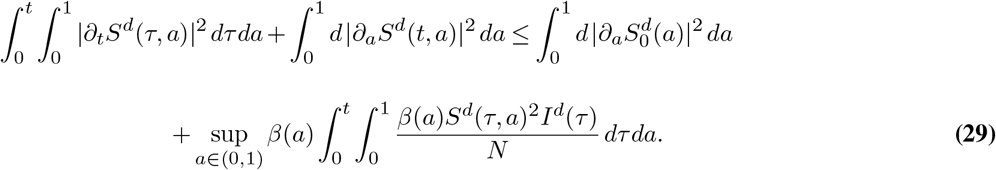

It follows that *∂*_*a*_*S*^*d*^ (*t*, ·) converges to 0 in *L*^*2*^ (0, 1): if it does not hold, there exists (*t*_*n*_)_*n*∈ℕ_ → +∞ and *α >* 0 such that 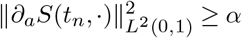. On the other hand, there exists *T* such that

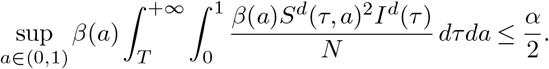

From (29) generalised between two positive times, for all *t > T*, there exists *n* such that *t*_*n*_ *> t* and

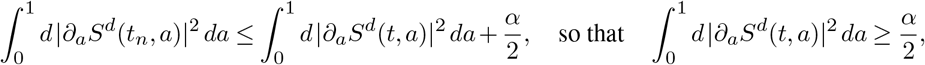

which contradicts the integrability of *∂*_*a*_*S* in *L*^2^ (ℝ^+^ × (0, 1)). Next, from (29), we deduce that *S*^*d*^ (*t*, ·) is bounded in *H*^1^(0, 1) uniformly in time, so that it is compact in *C*^0,1/2^(0, 1): a sequence (*t*_*n*_)_*n*∈ℕ_ → +∞ exists such that *S*^*d*^ (*t*_*n*_, ·) converges in *C*^0,1/2^(0, 1) to some 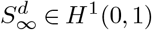 as *n* goes to +∞. Since 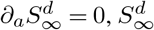 is a constant. In order to prove the uniqueness of 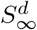, we observe that for all 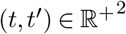

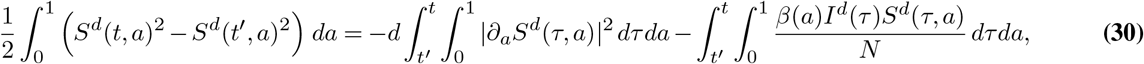

and use the fact that *∂*_*a*_*S* ∈ *L*^*2*^ (ℝ^+^ × (0, 1), *S/N* ≤ 1, and *I* ∈ *L*^1^ (ℝ^+^).

In order to prove (ii), using the fact that *dR*^*d*^ (*t*)*/dt* = *γI*^*d*^ (*t*) ≥ and *R*^*d*^(*t*) ≤ *N*, we deduce that *R*^*d*^(*t*) converges to some 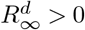. The conservation of total population 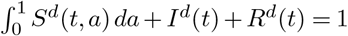 then yields the convergence of *I*^*d*^ (*t*) to 0 as *t*→ +∞.

Integrating (25) between 0 and *t*, and letting *t* go to +∞ leads to

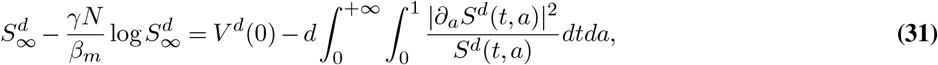

which yields the estimate 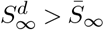. The detailed proofs and further developments are postponed to a forthcoming work.

### Some simulations with rebounds and plateaus

Some simulations show that the model adequately captures dynamical features such as epidemiological rebounds and plateaus. The initial conditions considered here are (18) where *κ* = 0.44, *S*_0_ = 90 and *S*_1_ = 119, 000 (total initial susceptible individuals are around 46, 398), initial infected people *I*(0) = 1, and *β* is given by (17) for *β* _0_ = 0.008 and *η* is taken such that *β*(1) = 26.

Diffusion of susceptible individuals seem to be the most sensitive parameter in order to obtain either traditional SIR type behaviour or rebounds and plateau-type dynamics. Figure 16 illustrates the changes of dynamics depending on diffusion parameter *d*. The associated dynamics of susceptible individuals are represented in Figure 17 when variable *a* ∈ (0, 1) is discretized into *n*_*g*_ = 50 groups through a finite difference scheme. Coloured curves represent each discretized group and the black curve the average of the groups (total susceptible individuals).

**Fig. 16.**
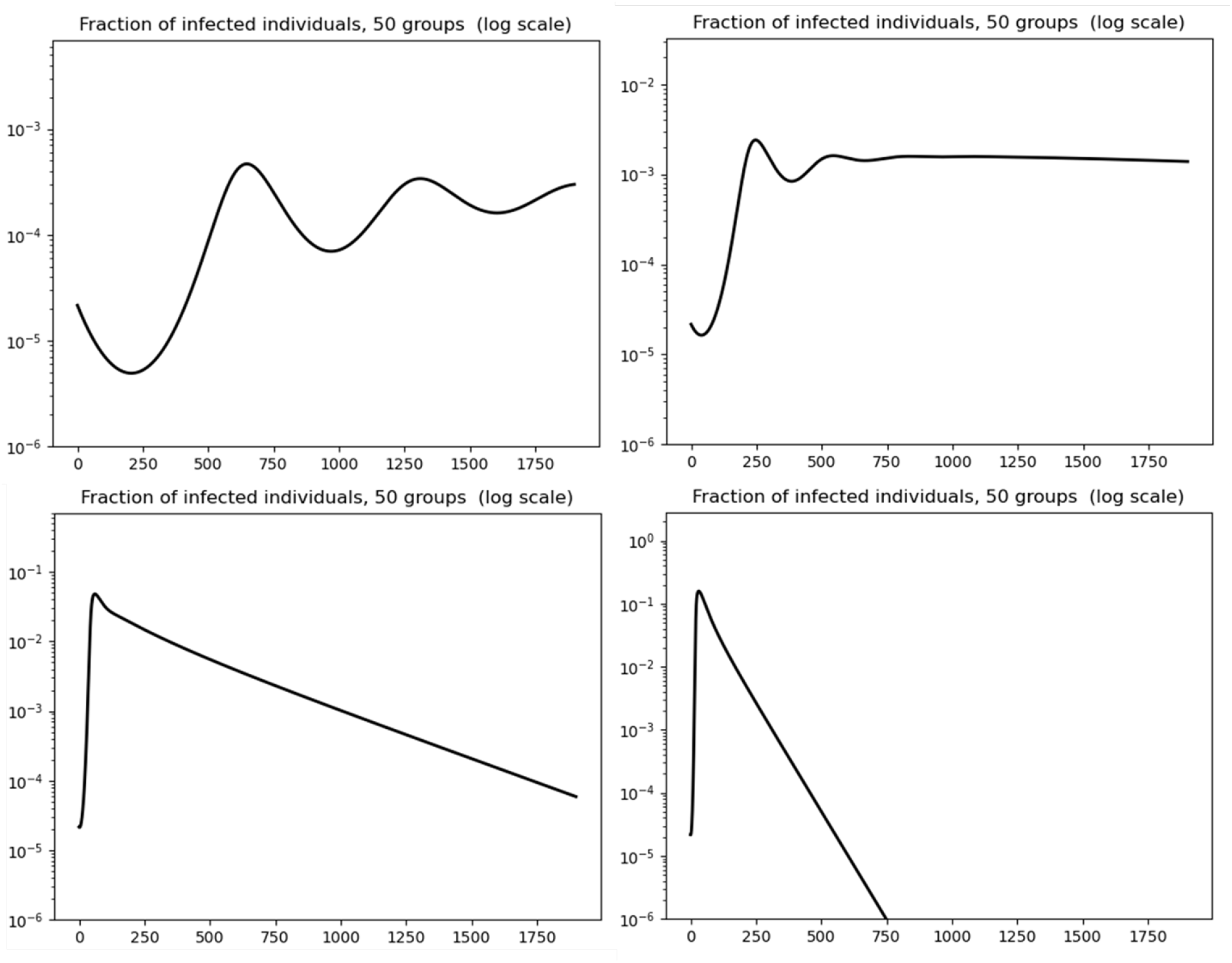
Model behaviour depending on diffusion parameter values: infected rate dynamics in logarithmic scale. From left to right and then top to bottom, graphs are associated with *d* = 10^−5^, *d* = 5 · 10^−5^, *d* = 10^−3^ and *d* = 5 · 10^−3^ (in *day*^−1^ unit).

**Fig. 17.**
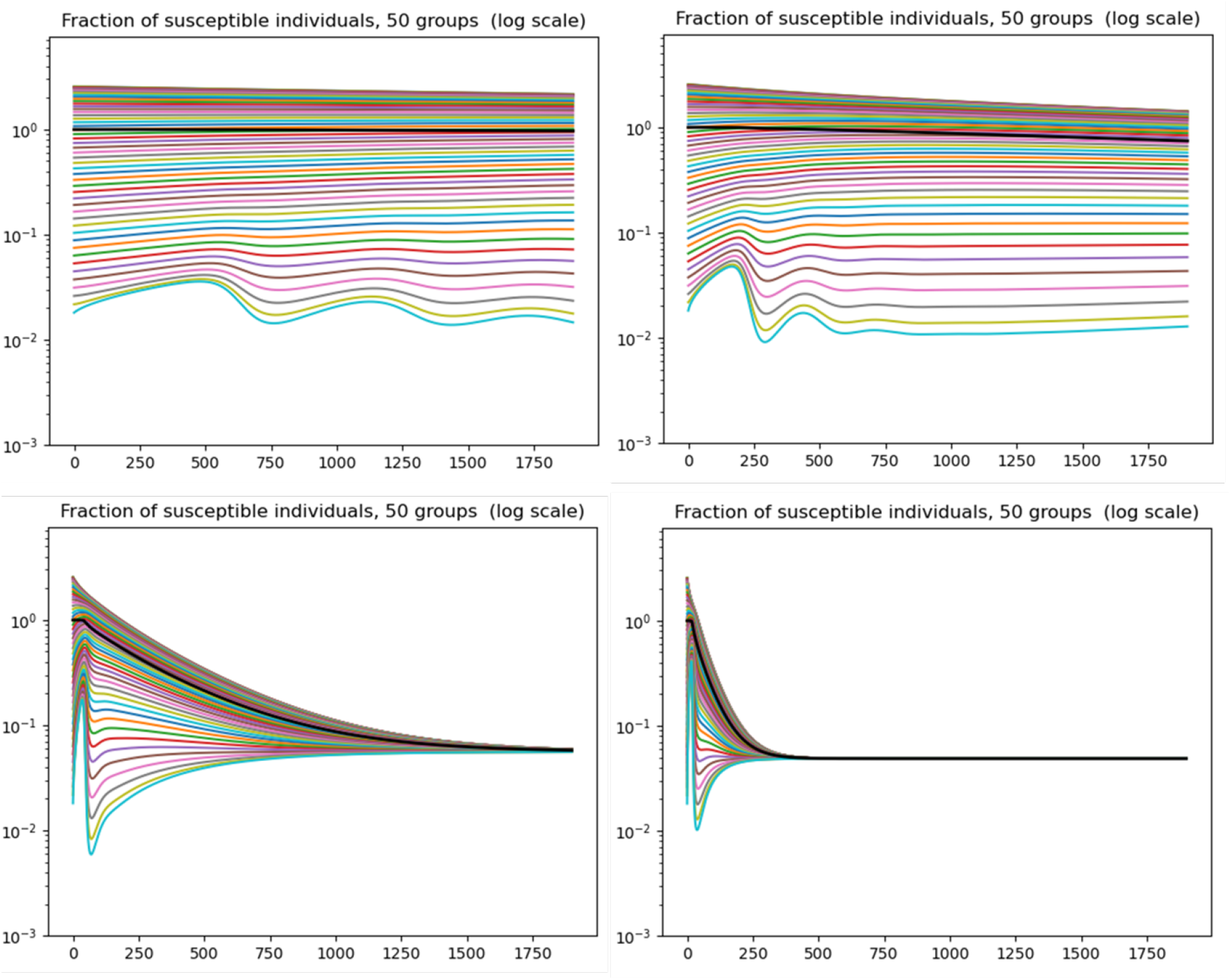
Model behaviour depending on diffusion parameter values: 50 groups of susceptible individuals, in logarithmic scale. The black curve corresponds to total susceptible individuals and the bottom light blue curve is associated with the most risky behaviour (*a* close to 1) and may not be monotonically decreasing in time. From left to right and then top to bottom, graphs are associated with *d* = 10^−5^, *d* = 5 · 10^−5^, *d* = 10^−3^ and *d* = 5 · 10^−3^ (in *day*^−1^ unit).

**Fig. 18.**
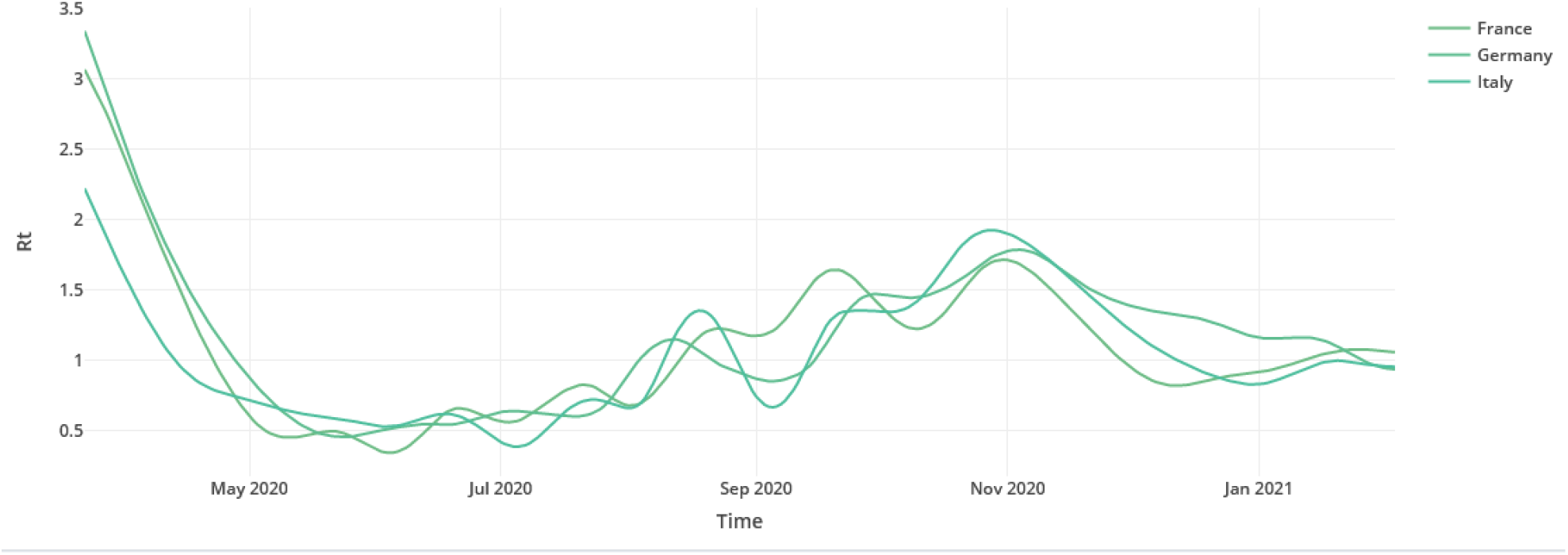
Effective *R* coefficient as a function of time (22) https://shiny.biosp.inrae.fr/app_direct/mapCovid19/.

One can observe the effect of diffusion: while individuals with the riskiest behaviour are rapidly consumed, their group is renewed through diffusion from the majority of less infectious individuals. Depending on the parameter range of the diffusion coefficient *d* in (12), it leads to multiple epidemic waves, plateaus, or classical single wave SIR-like dynamics.

### Stability of plateaus: the key role of diffusion

Let us illustrate in a heuristic manner the impact of diffusion on the existence and stability of plateaus in the dynamics of infectious individuals in Model (12). First, the second derivative of *x* = log *I* satisfies

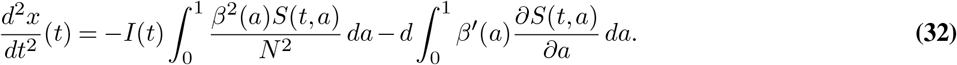

In the case *d* = 0, we deduce from (32) that *d*^2^*x/dt*^2^(*t*) *<* 0 for all *t* ∈ ℝ^+^, so that *x* = log *I* is a strictly concave function of time. The consequence is that only slow decrease at the beginning of the decay phase can be obtained. Plateaus or shoulder-like patterns cannot arise in the case *d* = 0, nor rebounds since *x*(*t*) has a single maximum after which it keeps decreasing.

In the case *d* ≠ 0, Equation (32) can be reformulated as

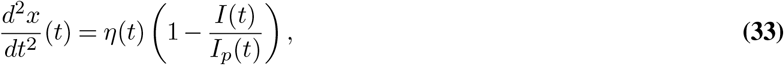

where

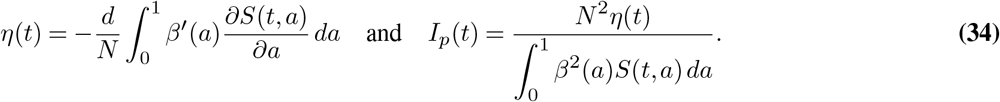

Assuming that *β* and *S*_0_ are respectively increasing and decreasing functions of *a*, we deduce from Proposition 1 that *η*(*t*) and *I*_*p*_(*t*) are always positive. Introducing *x*_*p*_(*t*) = log *I*_*p*_(*t*), Equation (33) can be rewritten as

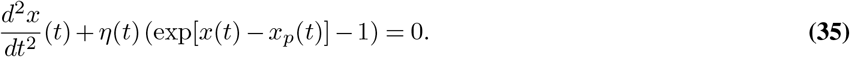

It means that *d*^2^*x/dt*^2^(*t*) may be positive (resp. negative) depending on whether *x*(*t*) is lower (resp. greater) than *x*_*p*_(*t*). In particular, in the neighborhood of times *t* such that *x*(*t*) is close to *x*_*p*_(*t*),

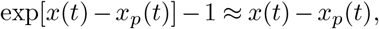

so that oscillations may arise, leading to patterns such as the ones seen on Figure 16. These oscillations also explain rebounds, or shoulders-like patterns before plateaus. Of course, this is not a proof but an indication to this effect and it warrants further mathematical investigation.

### More general systems

First, it should be noted that the coefficient *β*(*a*) can be time dependent: *β* = *β*(*t, a*). We can thus incorporate public health policies changes such as social distancing, lockdowns etc. For instance, we can consider a transition between two profiles *β*_1_(*a*) and *β*_2_(*a*) at a given date.

Then, we observe that System (12) is a particular case of a more general class of models. Indeed, we may want to also consider heterogeneity in the compartment of infected. This for instance reflects the division between symptomatic and asymptomatic individuals, or other differences. In particular, in terms of behaviour one can then include effects such as “super-spreaders”. We then get a model where the population of infected also depends on the same parameter *a*, that is *I* = *I*(*t, a*). We assume that the probability of interaction between a susceptible of class *a* and an infected of class *b* is given by a coefficient of transmission 𝔅(*a, b*). These considerations lead us to the following general system.

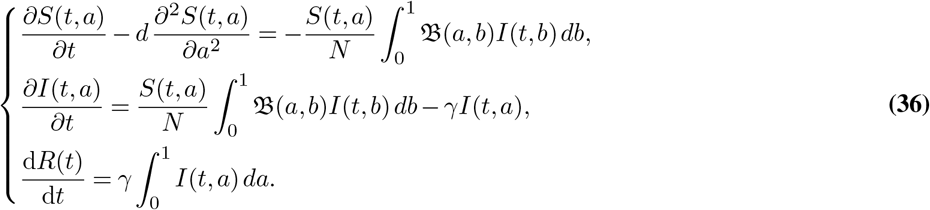

We observe that our model above (12) is derived from this more general class by assuming that 𝔅(*a, b*) is independent of *b*. In fact, the same is true for a finite number of states. This type of modelling of heterogeneity can be found in Magal et al.(16) and Arino et al. (13) in the discrete setting (*a* has a finite number of values) and without diffusion (*d* = 0). These works do not seem to yield shoulder, plateau and rebound-type patterns.

For simplicity we assume that the decay rate *γ* is uniform. Note that the second equation states that new infected individuals of class *a* result from an infection of a susceptible individual of the same class. Other choices are possible, but then, in order to be consistent, one would need to introduce new compartments such as symptomatic/asymptomatic, hospitalised etc. Likewise, we could consider other variants of the SIR model, such as the SEIR model. It would then be natural to assume that exposed individuals also move around. We are then led to a system of the following kind.

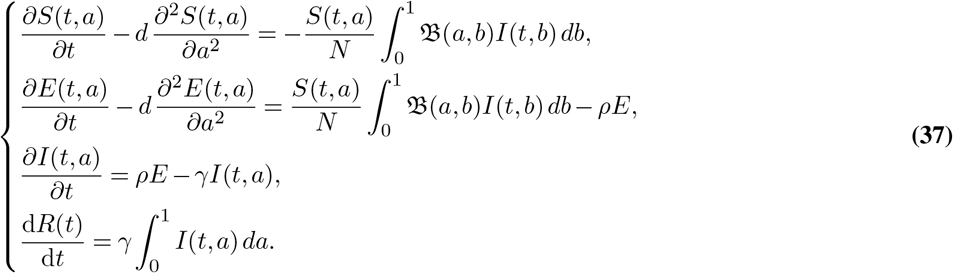

We can also consider a more general diffusion process on the behavioural variables than Brownian diffusion. We already mentioned above Eq. (11) the version involving a diffusion 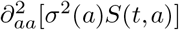 rather than 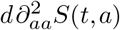.

With a somewhat different point of view, we can assume that there is a probability kernel *K*(*a, b*) that represents the probability distribution that an individual with trait *b* jumps to behaviour *a*. Then, the variation of *S* involves an integral operator rather than a heat operator. We are then led to the following system.

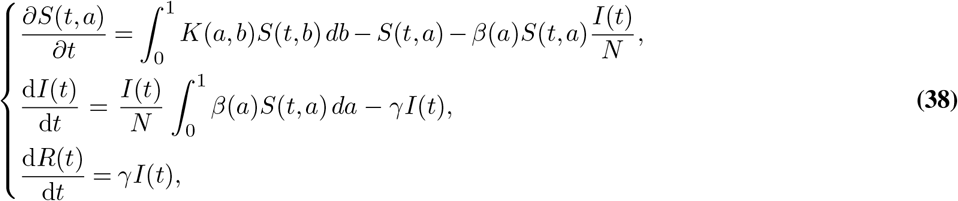

System (38) is supplemented with the same type of initial conditions as above (12). Note that there are no boundary conditions in this case.

It is classical that one can recover System (12) as a certain limit of the more general class of systems (38) (compare e.g. the article by the first author et al. (40)). However, here it is interesting to look at more behavioural assumptions on the kernel *K*. For instance, it is natural to assume that *K*(*a, b*) is singular when *b* is close to *a*, that *K*(*a, b*) is rather large when *b* is close to 1, whereas it is very small when *b* is small. We leave the study of such kernels for future study.

### Digital PCR in the framework of wastewater based epidemiology

Wastewater-based surveillance has been used over the past two decades to assess in real time the exposure of people to chemicals (in particular pollutants, licit and illicit or misused therapeutic or other drugs) and pathogens (bacteria or viruses) at the community level (41). It provides a global picture of population health at the scale of an urban area connected to wastewater treatment plants (WWTP). Fully complying with the privacy of individuals, this approach rests on an explicit, objective observation. It also has the advantage to serve as an early warning system. Nonetheless, there are also uncertainties in the quantitative interpretation of these measurements. In particular, dilution due to storm-water, infiltration of parasite inflow water, varying population, bio-marker degradation, varying time spent in the flow…, all generate a variety of biases.

In spite of these uncertainties, there is a renewed interest in wastewater based epidemiology (WBE), in the context of the Covid-19 outbreak. There are numerous initiatives over the world, in Australia (27), Japan (28), Netherlands (29), Detroit (30), Paris (31), Spain (32) and Canada (33). The recent periodic sampling campaigns in WWTPs rely on classical quantitative real time PCR (qRT-PCR) to estimate the concentration of SARS-Cov-2. At this stage, these programs serve as early warning platforms based on the observation that they detect SARS-Cov-2 in sewers at least seven days ahead of individual testings (30, 32, 34).

Only a few of them also used digital PCR on sub-samples to assess the measurement uncertainties of qRT-PCR measurements. Even if the benefit of dPCR in particular for low viral loads is mentioned (33), it is not currently used routinely for systematic measurements, mainly for economic reasons.

### Plateau-type patterns at country level

In most European countries, Covid-19 data exhibit similar plateau-type features, as illustrated in the so called effective *R* coefficient computed from the daily number of deaths (25). This effective *R* coefficient exhibits phases where it stabilises around 1, which corresponds to an epidemic plateau.

Campo et al. (42) study more specifically the dynamics of Covid-19 in Italy and put emphasis on plateaus and multi-wave patterns, for which they discuss meta-stability properties.

Press articles illustrate the political leaders’ observation that the Covid-19 outbreak has reached a plateau phase: Reuters, January 21^*st*^, 2021 (43); French government information service, December 10^*th*^, 2020 (44); A. Merkel, Westdeutsche Zeitung, January 21^*st*^, 2021 (45); E. Macron, February 2^*nd*^, 2021, interview on TF1 TV channel (46)).

